# The polygenic risk score and inter-familial heterogeneity in multigenerational families affected by schizophrenia and bipolar disorder

**DOI:** 10.64898/2026.06.08.26354912

**Authors:** Jasmin Ricard, Alix Dubeau, Claudia Moreau, Marie-Claude Boisvert, Michel Maziade, Alexandre Bureau, Simon L. Girard

## Abstract

In the past two decades, the focus on genome-wide association studies in large samples of unrelated patients has overshadowed family genetic studies. Therefore, little is still known about the levels and effects of the transmission of polygenic risk scores (PRS) among familial cases of schizophrenia (SZ) or bipolar disorder (BD) and their unaffected relatives. Prior research has shown that PRS are elevated in both patients and young individuals at familial risk for BD and SZ. We sought to study the transmission of PRS in affected multigenerational families and non-affected adult relatives (NAARs) with or without other non-mood nonpsychotic DSM-IV diagnoses and unrelated non-affected individuals from the same population. We genotyped 1,117 participants divided in 48 families from the Eastern Quebec Schizophrenia and Bipolar Disorder Kindreds. PRSs for both SZ and BD were computed using Multivariate Lassosum. For both SZ PRS and BD PRS, SZ and BD cases present higher PRS compared to controls, replicating previous findings. Regardless of a diagnosis of other non-psychotic and non-mood conditions, NAARs presented higher PRS than the unrelated cohort. Crucially, a subset of families presented consistently low PRS transmission profiles across generations, falling below expectations from our polygenic inheritance model. When the effect of individual PRs is accounted for, we observed sex-specific associations between familial PRS and patients’ symptom dimensions. Our results clearly demonstrate that polygenic inheritance alone does not adequately explain disease transmission in families. Such an approach may also clarify why some families exhibit dense clustering of cases despite minimal polygenic burden.

## Introduction

Psychiatric disorders such as schizophrenia (SZ) and bipolar disorder (BD) are complex and heterogeneous diseases for which the exact pathophysiology remains unknown. In this regard, there has been a large emphasis on genome-wide association studies (GWAS) conducted in the past two decades in large samples of unrelated patients and controls. Notwithstanding an important contribution, GWAS have provided a partial portrait of the genetic architecture of SZ and BD (1). An important GWAS contribution is that it confirmed that these disorders have a high heritability (2, 3) and a polygenic basis (4, 5). The millions of common single nucleotide polymorphisms (SNPs) with small effects identified in GWAS have contributed to the surge of subsequent studies of polygenic scores (PRS) for SZ and BD in several populations.

PRS aggregates the effects of common genetic variants to estimate an individual’s genetic liability to a given disorder. Despite its notable theoretical utility, PRS currently explain only a moderate portion of the SZ and BD liability variance (6) which limits its potential clinic applicability. Current estimates suggest that PRS account for approximately 11% of the liability variance in SZ (7) and 4% in BD (8). Also of note, there is evidence that the PRS is associated with parental history and specific characteristics of BD and SZ phenotypes in both non-familial and familial major psychiatric disorders. (9-11). This aligns with a renewed interest in family-based studies (12, 13). Our prior research (14), along with others, has shown that PRS was elevated not only in patients and their unaffected relatives but also in youths at familial risk for BD (11, 15, 16) and SZ (17, 18).

An important consideration is that the etiologic heterogeneity of SZ and BD (19, 20) may be one of the major obstacles to rapid progress in the genetic understanding of the disease, and PRS may help shed light on the heterogeneity of such complex disorders. Nevertheless, little is known regarding how the PRS may affect the transmission of SZ or BD among highly familial cases or in sporadic cases (21). Additionally, investigating sex-specific associations within familial contexts may reveal whether genetic risk is differentially expressed across sexes (22, 23).

Beyond case-control discrimination, an important question is whether polygenic risk scores predict clinical heterogeneity within affected individuals. Most studies have examined PRS at the level of the individual either at risk or affected by the disorder. To our knowledge, no studies have so far addressed the role and genetic effect of the whole familial PRS level on individual affected family members such as the effect of averaged PRS or other familial patterns of familial PRS.

Based on these previous observations, *our hypotheses* were: i) that PRS would distinguish the highly familial patients but also, their non-affected adult relatives (NAARs) from population controls and, ii) that PRS would not display the same transmission patterns among all multigenerational families, thereby allowing the identification of different subgroups of families characterized by their intra-familial PRS profiles.

In a large sample of 48 extensively phenotyped multigenerational families of Eastern Quebec (19, 24), our primary objective was to characterize the architecture of the PRS profile across the generations and assess whether fine-scale transmission patterns can differentiate between these large multi-affected families. We also sought to investigate sex-specific associations between familial PRS and clinical phenotypes. A necessary step was to confirm in our founder population the presence of a higher level of PRS in these highly familial patients having SZ or BD, and in their non-affective adult relatives, as previously found in GWAS extensive samples of unrelated mostly non-familial patients or relatives.

## Data and Methods

### Sample and Diagnosis

Families: We analyzed 1,117 individuals from the Eastern Quebec Schizophrenia and Bipolar Disorder Kindreds which comprises multigenerational families affected by SZ and BD(19, 25). The participants were from 48 multigenerational families recruited in the regions of Beauce, Saguenay-Lac-St-Jean, and New Brunswick-Îles-de-la-Madeleine in Canada. Given the length of time to recruit such an extensive sample, we made lifetime DSM-III-R or DSM-IV diagnoses that were based on best-estimate procedure and multiple sources of information and made by four psychiatrists who blindly reviewed all available lifetime information from different sources (26, 27). The narrow SZ definition was restricted to SZ, and its broad definition comprised SZ narrow plus schizophreniform disorder and schizotypal personality. The BD narrow phenotype was restricted to BD-I and its broad definition included BD-I, BD-II and recurrent major depression. We also defined a narrow and a broad common phenotype. The narrow common phenotype definition included BD narrow, SZ narrow and schizoaffective disorder (SAD). The broad common phenotype included the broad definitions of BD, of SZ, in addition to SAD. The specific numbers of cases for the narrow and broad phenotypes are found in Table 1. The details of the assessments of Global functioning with the Global Assessment Scale (27), age of disorder onset and the patient’s symptoms dimensions with the 82 items of the Comprehensive Assessment of Symptoms and History (CASH) (28) are provided in Supplemental Methods 1.4, Supplementary Table 1 and Supplementary Figure 1.

**Table 1:**
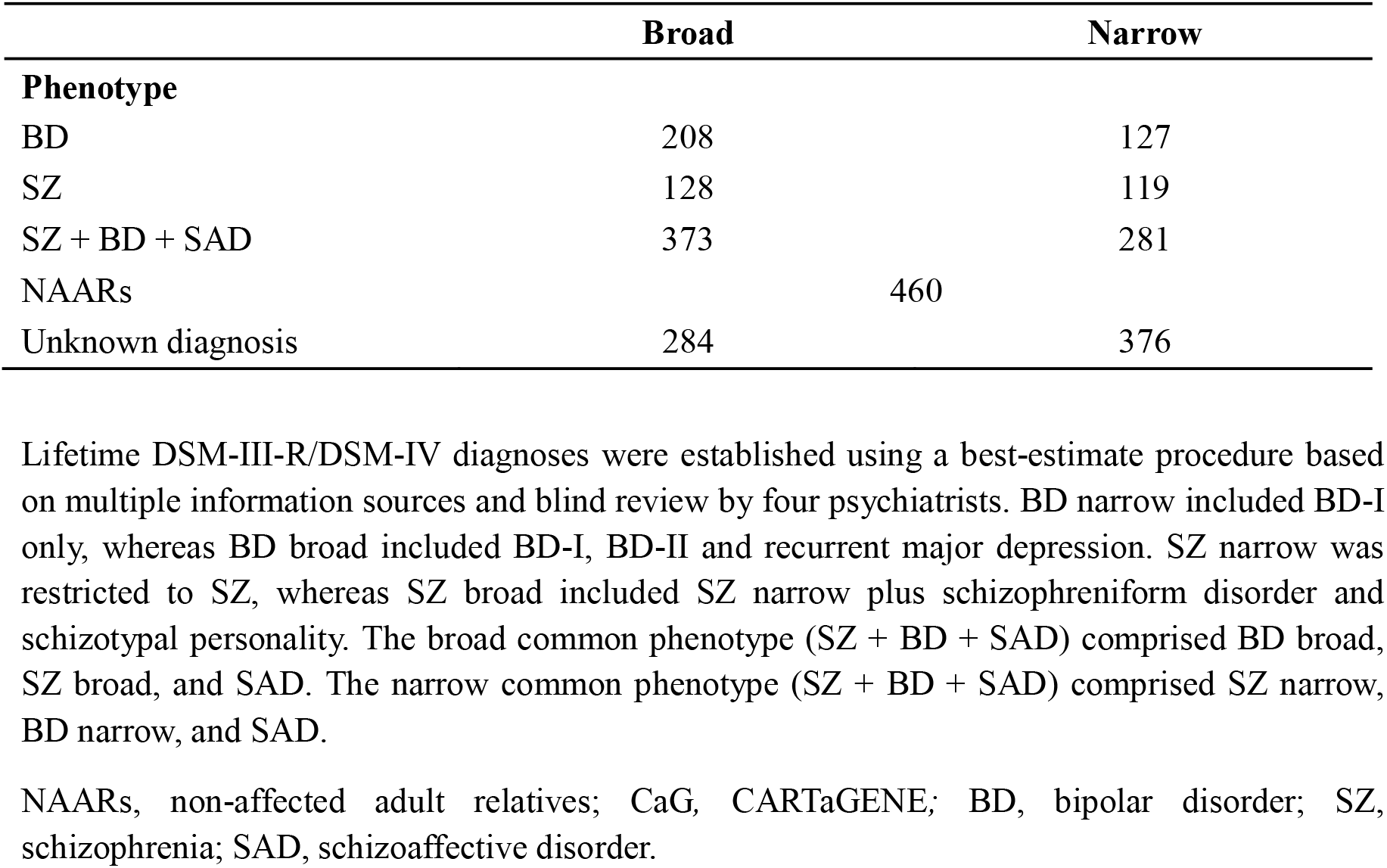
Phenotypes of the 1,117 Eastern Quebec Schizophrenia and Bipolar Disorder Kindreds cohort sample.

NAARs were subdivided into two categories according to the presence or absence of other non-mood, and nonpsychotic DSM-III-R and DSM-IV diagnoses (detailed in Supplementary Methods 1.1). Many NAARs (n = 140) presented at least one of the former diagnoses while the remaining 320 presented no such diagnosis (Supplementary Table 2). This study was approved by the Research Ethics Committee of the Centre Intégré Universitaire de Santé et de Services Sociaux de la Capitale-Nationale, Neurosciences and Mental Health Sector (Quebec, Canada). The reviewed and approved protocols included: *Genetic and Phenotypic Heterogeneity of Multigenerational Pedigrees Affected by Schizophrenia or Bipolar Disorder in Eastern Quebec (SPAP)* and *Neurocognitive Markers and Identification of Susceptibility Genes for Schizophrenia and Bipolar Disorder (Downtop)*. The study was conducted in accordance with ethical principles and institutional requirements applicable to research involving human participants. Written informed consent was obtained from all participants prior to their inclusion in the study. For participants under 18 years of age, written informed consent was obtained from a parent or legal guardian, in accordance with the requirements approved by the Research Ethics Committee.

The CARTaGENE (CaG) cohort provided control subjects (29). It included 2,184 whole genome sequences (WGS) from randomly selected individuals residing in both metropolitan and regional areas of the province of Quebec, Canada. To match the ancestry of the family members under study, only CaG participants of confirmed Quebec origin were retained, resulting in a final sample 1,884 individuals.

### Genotyping, Whole Genome Sequencing and Quality Control

SNP array genotyping was performed for the 1,117 related subjects using DNA extracted from immortalized lymphocytes or fresh blood by affinity column (Midi prep Qiagen). In a first wave 506 subjects were genotyped at 651,692 autosomal variants with the Illumina Infinium Human OmniExpress array. In a second wave, the remaining 611 subjects were genotyped for 691,719 variants with the Illumina Global Screening Array (GSA). Part of this sample was included in our previous studies (30, 31). Genotyping data were lifted over from the GRCh37 to the GRCh38 build using the LiftOver Genome Browser software (32).

On 464 related subjects, WGS was performed at the CHUL. DNA was extracted from stored frozen blood leukocytes and was prepared using the TruSeq DNA PCR-free library. Among them, 433 were also genotyped, while the remaining 31 subjects were only sequenced. The DRAGEN Germline Pipeline v3.10.4 on the Illumina BaseSpace was used to align the FASTQ on the hg38 Human reference genome and to call variants. These WGS were used to impute whole-genome genotypes using both population- and family-based imputations software.

For the CaG cohort, sequencing was done following the standard Illumina protocol. Further details on CaG WGS are available in this documentation (https://cartagene.qc.ca/files/documents/other/Info_GeneticData3juillet2023.pdf). A joint variant calling was conducted between the WGS gVCF files from both cohorts using the DRAGEN PopGen Joint Genotyping workflow via the Population explorer WebApp V4.0 (32). This produced a single combined VCF file of 79,000,100 positions (chr 1 to X) for 2,348 subjects.

QC was performed on the resulting joint VCF using PLINK 1.9b software (33). We excluded variants with more than 10% of missing calls, those with an allele coded as “NON_REF”, and monomorphic variants. Variants found in the centromeres and the ENCODE blacklist (34) were removed and variants found in the 1000 Genomes GRCh38 genome accessibility mask (35) were kept. Supplementary Figure 2 presents a detailed flowchart of the number of variants removed at each step. After QC, 36,096,233 autosomal variants remained. Genotypes involved in Mendelian errors were set to missing, prior to the imputation described below (34). Details about the genotyping data QC can be found in Supplementary Methods 1.2. We retained only variants that were also present in the QCed WGS, which left us with respectively 470,161 and 600,720 variants for the GSA array and the OmniExpress array. Supplementary Table 3 summarizes the number of variants and subjects from each source.

### Phasing and Imputation

Imputation was performed for the 684 subjects without WGS data. We first phased both WGS and genotyping datasets using ShapeIt5 (36). We then used two types of imputation: 1) family-based with GIGI2 (37) to leverage family relationships between our subjects and to improve the accuracy of rare variants imputation, 2) population-based with IMPUTE5 (38) allowing us to use 464 WGS and 36 additional unrelated subjects’ WGS, but also the 1,884 CARTaGENE sequences as an accurate ancestry-matched reference panel and to achieve better common variants imputation precision (see Supplementary Figure 3) (39). Finally, both imputations were combined by selecting the imputed genotype with the highest variance in its estimated posterior probabilities (40). Additional details are provided in Supplementary Method 1.3.

### Polygenic Scores

PRS were computed using the most recent BD (41) and SZ (42) summary statistics using the MultivariateLassosum (mvL) software (30). This method implements a penalized regression framework, leveraging the well-established genetic correlation between both traits (43, 44) to achieve a better prediction. We adopted a pseudo summary statistics approach, as proposed by (44), adapted to a multivariate scenario as detailed in (45) to select the best lambda parameter in the model without having to separate our dataset in training and validation sets. This method needs an external reference panel to estimate an LD matrix, we exploited the 1000 Genomes Project samples 404 Non-Finnish European subjects (46).

Analyses were restricted on 5,189,652 autosomal SNPs in our WGS also found within all datasets. Then, we further restricted this number to 3,717,443 autosomal SNPs with a MAF > 5%. The S-LDXR software (47) was employed on SZ and BD summary statistics and the CaG sample genotypes to obtain LD score estimates. This result was then used within S-LDXR to estimate per-SNP contributions to the heritability and genetic covariance of both traits on: 1) pseudo summary statistics, 2) the real GWAS summary statistics. Finally, mvL was first launched on the pseudo summary statistics to select the best tuning parameters and it was launched a second time on the real GWAS summary statistics to estimate the actual BD and SZ PRSs.

### Statistical Methods

Analyses were conducted using R (48). Logistic regressions accounting for familial structures was performed through generalized estimation equation (GEE). This was achieved using the R package *geepack* V1.3.12 (47). PRS for BD and SZ were standardized (mean = 0, SD = 1). Depending on the analysis, controls were either CaG or NAARs (Supplementary Table 2). This methodology enables different odds ratios (ORs) for these disorders for an increase of 1 PRS standard deviation. We also estimated areas under the receiver operating curve (AUC). PRSs were dichotomized for further analyses. To do so, we used the reference group to find the 75% PRS percentiles for men and women, then, subjects presenting a PRS higher than these thresholds were classified as “higher PRS” group.

When looking at the degree of heterogeneity among families in their profiles of PRS transmission, we identified families with more homogeneous PRS among their affected members and among their NAARs (in other terms, families with a limited within-family variability) before examining whether their affected members have higher or lower PRS than expected. We computed mean PRS across all affected members to create a familial PRS and the pooled standard deviation of PRS for patients (broad SZ + BD + SAD phenotype) and NAARs. Focusing on families with at least 3 SZ or BD cases and low pooled SD, we evaluated potential heterogeneity between families in their mean PRS level among cases, highlighting families where the PRS was homogeneously low (mean < 0), high (mean > 1) or intermediate. We then evaluated the probability of observing lower SD values as well as lower PRS than the maximum PRS observed in low PRS families, and conversely PRS higher than the minimum PRS observed in high PRS families. Since we are not aware of existing models of PRS in large extended families accounting for the number of affected and unaffected members and their relationships, we formulated a simplified model and a simulation approach to estimate probabilities (described in Supplementary Methods 1.4).

Given our results on familial PRS, we attempted an external clinical validation of the result by examining the potential associations between the familial PRS and phenotype i.e., age of disorder onset, global functioning and symptom dimensions. Assessment procedures for these clinical variables and the derivation and interpretation of symptom components through PCA are detailed in Supplementary Methods 1.5. Analyses used linear mixed-effects models (LMMs) with random family intercepts. Individual and familial PRS (across all affected members) were included in the models to separate individual-level and cluster-level effects (49). In addition to marginal PRS effects, sex-PRS interactions were tested, and sex-stratified effects were estimated. Families were weighted by the inverse of the PRS variance among their affected members to upweight more homogeneous families in terms of the patients’ PRSs. Multiple-testing correction used the Benjamini-Hochberg (BH) method to control false discovery rate (FDR) at 10% across 16 tests. BH-adjusted *p* values (*q* values) were computed using the multtest package in R.

## Results

Our results first confirmed that PRSs were significantly higher among patients compared to NAARs (PRS SZ OR for broad SZ cases vs. all NAARs = 1.83 [1.48–2.26], *p* = 1.9×10^−8^; PRS BD OR for broad BD vs. all NAARs = 2.1 [1.71–2.58], *p* = 5.0×10^−11^). The difference remained significant regardless of the presence or absence of other non-SZ and non-affective diagnoses (PRS SZ OR with vs. without other diagnosis = 0.97 [0.77– 1.21], *p* = 0.18; PRS BD OR with vs. without other diagnosis = 1.15 [0.93–1.42], *p* = 0.2). Furthermore, when compared to controls, PRSs were higher in NAARs, especially for SZ NAARs (NAARs without diagnosis vs CaG, PRS SZ OR = 1.59 [1.39–1.83], *p* = 3.8×10^−11^; PRS BD OR = 1.21 [1.03–1.42], *p* = 0.026).

When assessing the specificity of the SZ PRS to the SZ phenotypes, stronger associations were observed with the narrow SZ phenotype than with the narrow BD, and also stronger than with the common phenotypes (Figure 1). Conversely, the BD PRS showed a stronger association with the narrow BD than with the narrow SZ. Predictive performance, as measured by the AUC, was consistent with the association strength measured by the ORs. As shown in Figure 1, these specificity patterns were consistent across the broad and the narrow phenotypes. Given our reported findings of genetic heterogeneity among different Québec populations (50, 51), we found here no significant differences among the Quebec province regions of recruitment (Supplementary Figures 4 and 5). We also stratified the result by sex and PRSs discrimination power remained the same among men and women (Supplementary Figure 6).

**Figure 1:**
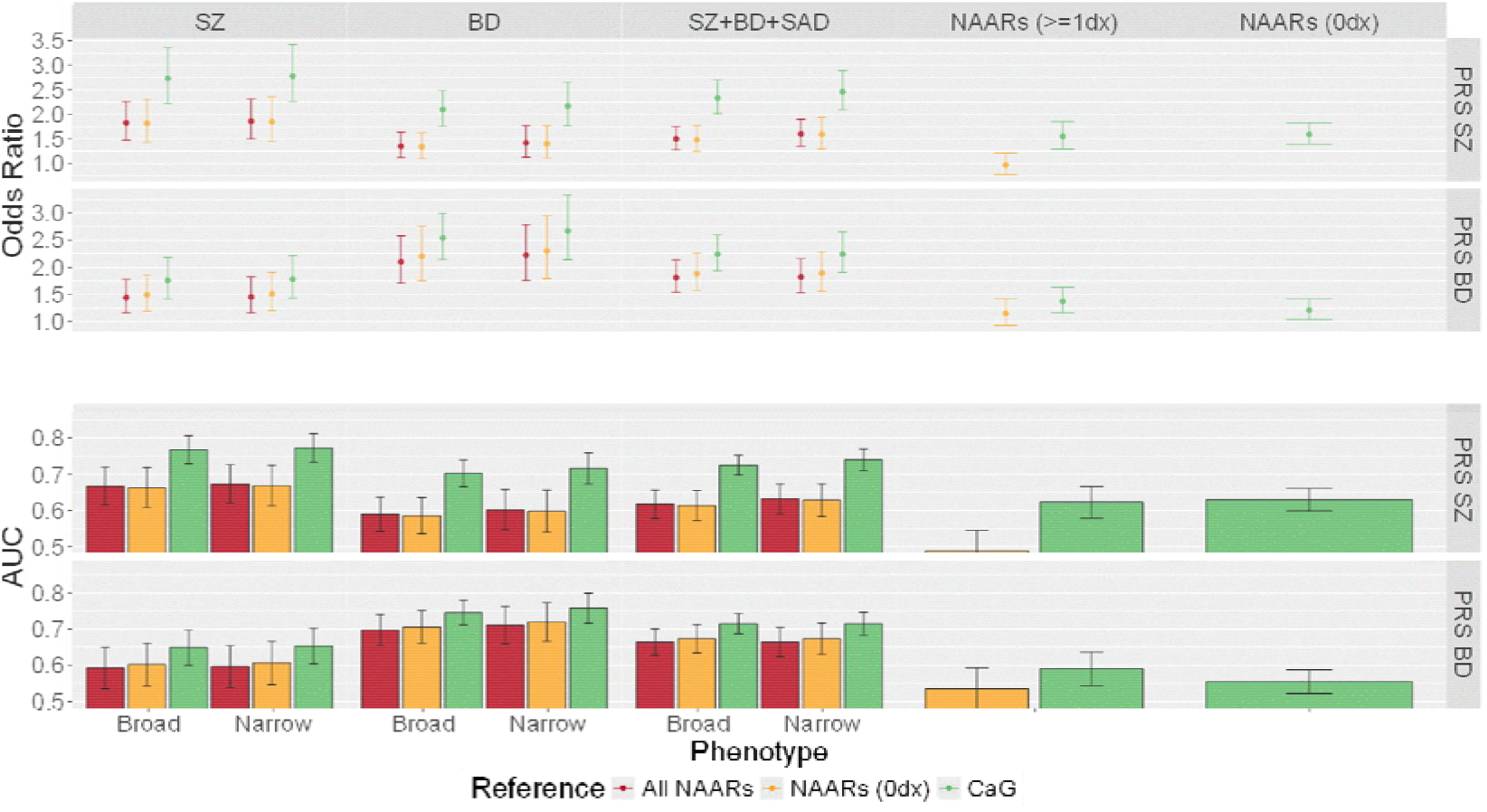
The predictive performance of the continuous SZ and BD PRS in distinguishing SZ, BD and SZ + BD + SAD cases – in their broad and narrow definitions – from various reference groups. Top panel: Odds ratio (OR) and 95% confidence interval of phenotype for an increase of 1 standard deviation in PRS. Bottom panel: Area under the receiver operating curve (AUC) and 95% confidence interval for the prediction of the phenotypes by PRS. Numbers of subjects used in each OR estimation are presented in Supplementary Table 2. NAARs (0 dx) refers to non-affected adult relatives without other non-mood non psychotic DSM-III-R or DSM-IV diagnosis. NAARs (≥ 1dx) refers to non-affected adult relatives presenting at least one other non-mood non psychotic DSM-III-R or DSM-IV diagnoses. BD narrow includes BD-I only, whereas BD broad includes BD-I, BD-II and recurrent major depression. SZ narrow is restricted to SZ, whereas SZ broad includes SZ narrow plus schizophreniform disorder and schizotypal personality. The broad common phenotype (SZ + BD + SAD) comprises SZ broad, BD broad, and SAD. The narrow common phenotype (SZ + BD + SAD) comprises SZ narrow, BD narrow, and SAD. PRS, polygenic risk score; NAARs, non-affected adult relatives; CaG, CARTaGENE; BD, bipolar disorder; SZ, schizophrenia; SAD, schizoaffective disorder.

Distinct transmission patterns were observed when examining the PRS across each multigenerational family. Within some individual families, PRS exhibited higher variability among their affected family members whereas other families had more homogeneous PRS among affected members (Figure 2). This observation prompted us to further investigate families where the PRS remained stable among relatives–specifically families that would be characterized by higher or lower overall PRS. Such families were selected because they displayed the lowest pooled PRS SD between cases and NAARs while exhibiting a low PRS (Figure 2A) or a high PRS (Figure 2B) among cases (BD PRS: 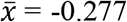 (SD = 0.541) and 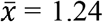 (SD = 0.492) as presented in Supplementary Table 5; SZ PRS: 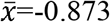 (SD = 0.577) and 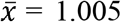 (SD = 0.372) as presented in Supplementary Table 6). The SD of each family for both PRSs against the total number of NAARs and cases are presented in Supplementary Figure 7. The observed low SD values of the selected families fall within the distribution of SD expected for these families under the simplified PRS model that we adopted (probability of SD lower than the observed values between 0.09 and 0.68, Supplementary Tables 7 and 8).

**Figure 2:**
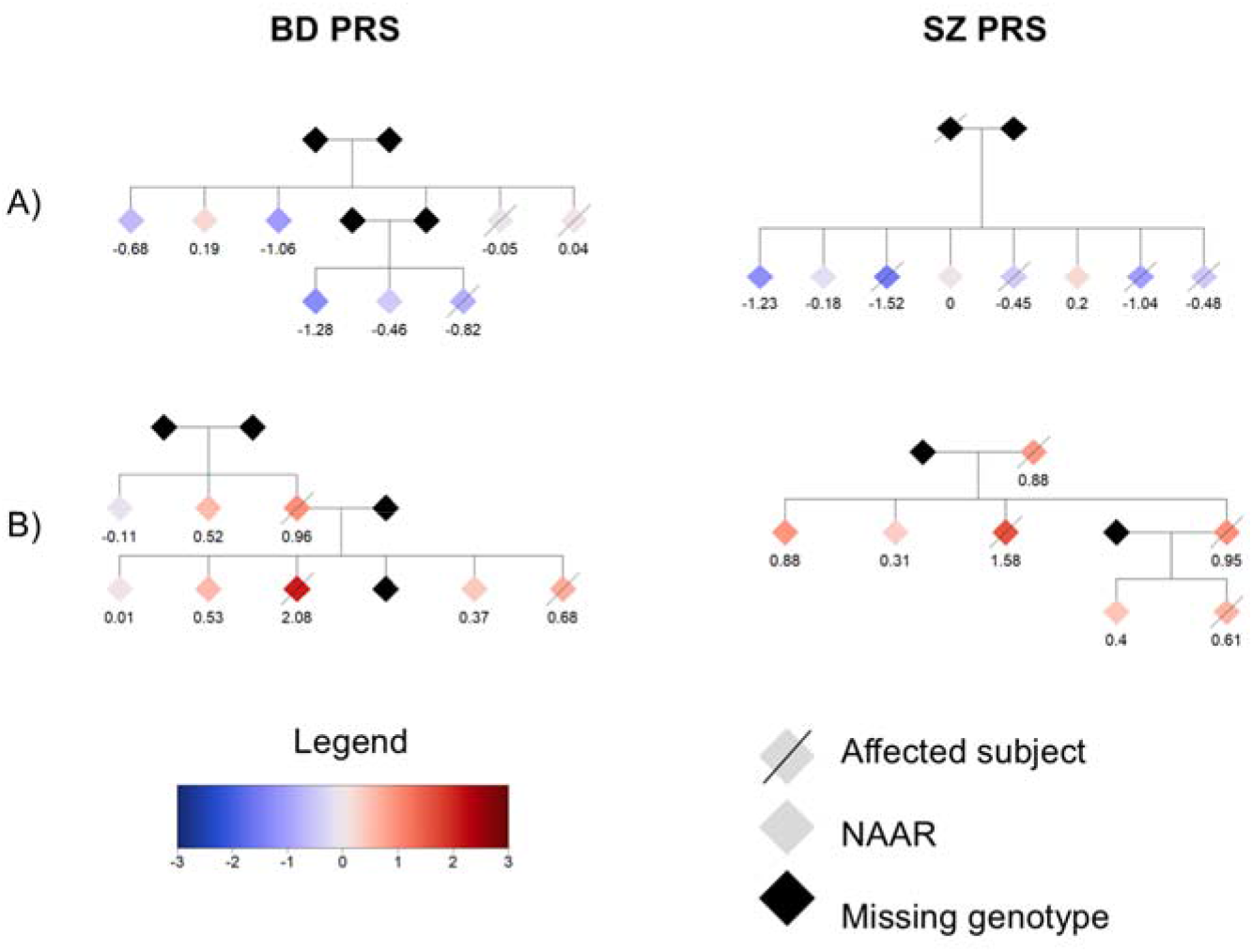
Examples of families that illustrate differences in overall SZ PRS and BD PRS levels and transmission. Affected subject refers to the broad SZ + BD + SAD phenotype, which comprises SZ broad, BD broad, and SAD. BD and SZ PRS were both scaled using the mean and the SD values among the CaG sample. The two families in A) would contain patients with SZ, BD or SAD as well as their NAARs having PRS in the lower end of the PRS distribution as explained in legend. The two families in PRS B) would contain patients with SZ, BD or SAD as well as their NAARs having PRS in the upper end of the PRS distribution PRS. Mean (SD) values were as follows: A) BD PRS, -0.277 (0.541); SZ PRS, -0.873 (0.577); B) BD PRS, 1.24 (0.492); SZ PRS, 1.005 (0.372) (Supplementary Tables 6 and 7). To preserve the anonymity of the patients, biological sex as well as several individuals were removed from the graphical representation. PRS, polygenic risk score; NAARs, non-affected adult relatives; CaG, CARTaGENE; BD, bipolar disorder; SZ, schizophrenia; SAD, schizoaffective disorder; SD, standard deviation.

However, the probability of observing PRS values lower than the maximum observed for the selected families with low PRS values was small under the same model (p < 0.03 for the cases + NAARs, *p* < 0.07 for the cases only under the most conservative RR value of 1.3. Supplementary Table 7). In contrast, the probability of observing PRS values higher than the minimum observed among cases in the selected families with high PRS values were large (*p* > 0.12 for the same RR = 1.3, Supplementary Table 8).

Based on our findings that families would differ in terms of their level of PRS transmission, we tested whether the familial PRS was associated with patients’ phenotypic characteristics (Supplementary Methods 1.5, Supplementary Table 9). First, the linear mixed-effects models (Supplementary Table 10) revealed no significant effect of familial SZ or BD PRS on age of onset and global functioning (GAS), with *q* values being > 0.60. The results were different regarding the patients’ symptom components as defined by the PCA applied to the CASH symptoms. We indeed observed a significant sex-by-PRS interaction for the familial BD PRS, a higher familial BD PRS being associated with a higher *mania* component score in males (*ME*_*M*_ = 0.57, 95% IC [0.21, 0.93]) but not in females (*ME*_*F*_ = -0.07, 95% IC [-0.43, 0.30], interaction *p* = 0.004, *q* = 0.064) (Supplementary Table 11, Supplementary Figure 8). We also observed a trend of association between the familial SZ PRS and the *mania* component, with higher familial SZ PRS associated with a lower *mania* component score in females (*ME*_*F*_ = -0.45, 95% CI [-0.88, -0.02]) but not in males (*ME*_*M*_ = 0.26, 95% CI [-0.23, 0.74]; interaction *p* = 0.016, *q* = 0.128) (Supplementary Table 11). Another trend was observed between the familial SZ PRS was associated with higher *schizophrenia* component score in males (*ME*_*M*_ = 0.42, 95% IC [-0.02, 0.90]) but not in females (*ME*_*F*_ = -0.16, 95% IC [-0.58, 0.27], interaction *p* = 0.040, *q* = 0.184) (Supplementary Table 11). Finally, no association was observed between familial BD or SZ PRS and the *depression* component in either males or females.

## Discussion

After having confirmed in our sample that the highly familial patients having SZ or BD and their NAARs had higher levels of PRS than control subjects as observed in previous studies, we observed a specificity of association between the SZ and BD PRS with their respective disorder; confirming that PRS captures a portion of the common genetic liability underlying these disorders. Interestingly, the discriminative power of the PRS did not increase when using narrower diagnostic definitions. Beyond a degree of specificity, our data suggest that SZ and BD PRS provide comparable predictive power across a broad spectrum of psychotic and affective phenotypes, also supporting a shared polygenic background between these disorders.

The original contribution of this work lies in our observation that multigenerational and densely affected families don’t have similar PRS transmission patterns. While most multigenerational families exhibited PRS levels consistent with expectations, a subset maintained consistently low overall PRS values across generations. The consistently low values of the intrafamilial PRSs fell below what would be expected under our simplified model of polygenic inheritance. This strongly suggests that, in this subset of multiaffected families more than any others, PRS alone would not explain the disease transmission. For these families, disease would be rather driven by other factors—such as rare, highly penetrant variants, or shared environmental exposures that interact with genetic susceptibility. Integrating such alternative mechanisms may be essential to build more comprehensive models of psychiatric disease inheritance in a context of genetic heterogeneity. Such an approach may also clarify why some families exhibit dense clustering of cases despite minimal polygenic burden. In contrast, families with consistently higher PRS values largely fell within the expected distribution, implying that their aggregation of illness would be consistent with polygenic inheritance. Together, these findings would illustrate, at least partly, the genetic heterogeneity underlying SZ and BD in highly familial cases: some families would follow a polygenic inheritance model, while others likely involve distinct, non-polygenic mechanisms.

We also evaluated the potential clinical significance of our findings by examining the association between familial PRS and three clinically relevant phenotypes in the semiology of SZ, BD, and MDD after accounting for the individual PRS in the analysis. We found no association with two broader and widely used phenotypes: age at disorder onset and global functioning. However, a significant association was obtained with a finer dimensional phenotype derived from a PCA of CASH symptoms. This analysis yielded a strong structure consisting of three components, which incidentally corresponded to the original constructs of the instrument. In males, the familial BD PRS showed a significant association with the *mania* symptom component, which explained 17.9% of total PCA variance, and there was a trend towards association between the familial SZ PRS and the *schizophrenia* symptom component, which accounted for 40.9% of the total variance. In women, a trend of association was observed between the familial SZ PRS and the *mania* component: higher familial SZ PRS was linked to lower mania-component scores, which corresponded to more depressive rather than manic symptoms. Finally, no association was found with the third *depression* component (11.8% of total PCA variance).

These data on extended phenotypes provide important elements of external clinical validity of our present finding of a differential familial PRS transmission among multiaffected families. They suggest an effect of the SZ and BD familial PRS, beyond the effect of the individual PRS, on the clinical features of affected family members in the form of a severity and sex-specific impact on schizophrenia and mania symptoms. They also suggest a specific effect of SZ or BP familial PRS at the level of the schizophrenia or mania symptom components, respectively.

Another important result of our study is that non-affected adult relatives (NAARs) of highly familial patients exhibited higher PRSs than population controls, regardless of whether they have other DSM diagnoses than the broad SZ or BD diagnoses. This observation reinforces the idea that PRSs would capture a general familial liability to psychiatric illness that extends broadly beyond SZ and BD themselves as again clearly suggested by the large study (52). Our longitudinal clinical follow-up of all NAARs allowed us to confirm that none met criteria for SZ or BD even if they carried a detectable genetic load. This highlights the importance of incorporating the family context in the studies sampling methods when interpreting genetic risk and supports the value of using well-defined population controls in psychiatric cohorts.

Regarding limitations, our model of PRS transmission excluded indirect effects and shared environment and assumed a rare disease. These simplifications likely explain the smaller differences in mean PRS between cases and NAARs predicted by the model compared to the empirical differences (Supplementary Tables 6 and 7). This reduced difference increases the probability of consistently low PRS across all family members under the model. Finally, we further emphasize that due to the nature of our data, findings cannot be generalized to non-familial individuals, due to this shared environmental and genetic background.

Altogether, our results reaffirm the utility of PRS for quantifying shared genetic liability in highly familial SZ and BD, while also underscoring their limitations. The presence of multigenerational families with unexpectedly low PRSs suggests that different genetic architectures coexist within SZ and BD, with some families following a highly polygenic trajectory and others shaped by rare variants or environmental factors. This heterogeneity argues against a one-size-fits-all model of genetic risk and calls for integrative approaches combining polygenic and rare variant analyses in extended family designs.

## Supporting information

All supplementary material

## Data Availability

Scripts in R code for the analysis of schizophrenia and bipolar disorder are available on Github (https://github.com/abureau/RV_in_SZ_BD_kindreds). The data of the Eastern Quebec SZ and BD kindred study are in the process of being deposited on the Centre Quebecois de Donnees Genomiques (CQDG) and accession number will be made available in the final version of the manuscript. The data from the CARTaGENE project are available after approval of an access request submitted at https://www.cartagene.qc.ca/en/researchers/access-request.html. The data are not publicly available due to privacy and ethical restrictions.

## Acknowledgements

We are grateful to Yvon C. Chagnon and Lise St-Germain for managing our molecular biology laboratory, to the professional research assistants: Louise Bélanger, Linda René, Lisette Gagnon, Claudie Poirier, Nicole Leclerc, Julie Lamarche, Pierrette Boutin, Mélanie Mercier, Jordie Croteau, Alain Fournier and to the family members who participated in the Eastern Quebec SZ and BD kindred study, which was funded by a Canada Research Chair (\#950-200810) in psychiatric genetics of which Maziade is the Chair and by Canadian Institute of Health Research grants (\#MOP-74430, \#MOP-114988, \#PCG-155471 and \#PJT-175122). This project was also funded through another Canada Rearch Chair in genetics and genealogy attributed to Girard. This research project was funded by the New Frontiers in Research Fund, Exploration grant 2019-00752. The data management system was supported by the Canada Foundation for Innovation Leadership Opportunity Fund (grant 27592).

## Supplementary Information

Supplemental Information includes additional details on methods, tables and figures.

## Availability of Data and Materials

Scripts in R code for the analysis of schizophrenia and bipolar disorder are available on Github (https://github.com/abureau/RV_in_SZ_BD_kindreds). The data of the Eastern Quebec SZ and BD kindred study are in the process of being deposited on the Centre Québécois de Données Génomiques (CQDG) and accession number will be made available in the final version of the manuscript. The data from the CARTaGENE project are available after approval of an access request submitted at https://www.cartagene.qc.ca/en/researchers/access-request.html. The data are not publicly available due to privacy and ethical restrictions.

