## Supplementary material for "The polygenic risk score and inter-familial heterogeneity in multigenerational families affected by schizophrenia and bipolar disorder": All supplementary material

Table of content

[Supplementary Table 7 **–** Probability of observing low PRS (defined as a mean PRS among cases < 0) with the broad SZ + BD + SAD phenotype in families from Figure 2A.	13](#_Toc231375094)

### 1 Supplementary Methods

#### 1.1 NAARs DSM-III-R and DSM-IV diagnoses

Diagnoses were grouped in four different categories by research diagnosticians. We did not consider diagnoses rated as merely “possible”, nor the diagnoses of personality disorders. Only probable and definitive diagnoses were considered for this study.

1) Mood: major depression (situational, uncomplicated, episodic), adjustment disorder, dysthymia, cyclothymia (n = 99);
2) Psychosis: delusional disorder, psychosis not otherwise specified (n = 6);
3) Anxiety: generalized anxiety disorder, specific phobia, panic disorder, social phobia, agoraphobia, obsessive-compulsive disorder, anxiety disorder not otherwise specified (n = 39);
4) Consumption: drug abuse, substance dependence, substance abuse (n = 59).

Our discovery cohort was recruited several years ago and followed in the Eastern regions of Quebec over a 12-year period which explains why DSM-III-R and DSM-IV diagnoses have been made. The inclusion criteria were having a first-degree relative with a definite DSM-III-R or DSM-IV SZ or BD diagnosis. The procedure included the Structured Clinical Interview for DSM-III-R or DSM-IV Disorders (SCID), family interviews, and all available medical records throughout the individual's lifetime, following a previously established method with demonstrated reliability. Subsequently, to validate the diagnoses, a group of four researcher psychiatrists independently established a best estimate DSM-III-R or DSM-IV diagnosis across the lifetime, and the reliability of this process was also examined.

#### 1.2 Genotypes quality control

SNPs with the following quality problems were removed: missing call rate higher than 0.02, SNP mismatches with Haplotype reference consortium (HRC), SNPs which are indels, minor allele frequency difference with HRC > 0.2, palindromic SNP with frequency > 0.4, allele mismatch with HRC, duplicates, Hardy Weinberg equilibrium test *p* value lower than 1x10e-7.

From the first wave of SNP array genotyping using an Illumina Omni Express chip at Genome Québec, a total of 507 subjects were genotyped at 651,692 autosomal SNPs. A total of 50,972 SNPs were removed following quality control. For 237 positions, no matching was found when a LiftOver from GRCh37 to GRCh38 was done. From the second wave of SNP array genotyping using an Illumina Global Screening Array at Genome Québec, a total of 615 subjects were genotyped at 691,719 autosomal SNPs. A total of 188,854 SNPs were removed following quality control. For 15,156 positions, no matching was found when a LiftOver from GRCh37 to GRCh38 was done.

#### 1.3 Phasing and simulation

To phase our QCed WGS, we used ShapeIt5 software (1) which allowed us to phase common and rare variants separately, while accounting for familial relationships between subjects. This method starts by phasing common variant (using a minor allele frequency (MAF) threshold of 0.1%, as recommended) to generate scaffolds of highly confident haplotypes. These scaffolds are then used to phase the rare variants. To maximize parallelization, we phased on large chunks of 25 centimorgans. Genotyping data are phased using the common variants pipeline only.

The GIGI2 imputation software (2) takes advantage of the family relationships. It requires, as an input, the inheritance vectors across the genome generated by the gl_auto software from the MORGAN 3.4 (3). gl_auto only requires a small set of independent SNPs spaced by 0.5 centimorgans. This was achieved by implementing a MAF-based pruning with a threshold of 35%, yielding 21,843 SNPs. Because the number of SNPs used in this process is small, we combined both arrays and retained only the SNPs found in the intersection between positions. Default parameters were used, except that each SNP MAF was set to 0.5. 30,000 Monte Carlo iterations were performed, and we did 1000 burn-in iterations. GIGI2 was run separately on each family. A total of 36,079,183 positions were imputed across the 1,117 subjects found in the 48 families, including 409 subjects without genotypes or WGS.

IMPUTE5 (4) extends the IMPUTE method (3) by using a selection of haplotypes based on the Positional Burrows Wheeler Transform (5), making it faster, more accurate and more memory-efficient. Default parameters were used, and the imputation was performed separately for each genotyping array using the WGS data from both CaG and our cohort as the reference panel. We argue that it is more advantageous to perform the imputation on a smaller number of subjects genotyped per chip while retaining a larger number of variants. For both chips, 36,091,936 positions were imputed.

To combine the family–and the population-based imputations, we used the ped_pop software (5, 6) on 1,117 subjects and 36,079,183 positions imputed by both GIGI2 and IMPUTE5. This software requires imputed genotype probabilities for both methods and a performance score for the population-based method (INFO score by IMPUTE5). The ped_pop software produced combined imputed genotype probabilities that were then used to call genotypes (probability threshold of 0.9) (7). Imputation accuracy was assessed using 47 samples sequenced with another library generation method (linked-reads from 10X). We performed a principal component analysis (PCA) to examine the juxtaposition of the 10X and Illumina samples on the principal components. Supplementary Figure 3 suggests no batch effect and a good agreement between both technologies. Furthermore, using PLINK, we compared the imputations with their 10x sequences and we obtained error rates ranging from 0.00808 to 0.0153 (mean = 0.0114; SD = 0.00165), which is similar to what was observed in (8).

#### 1.4 Model and simulation approach for joint distribution of PRS in families

We used a simplified model of PRS transmission in families to assess the probability of PRS vectors being more extreme than observations in families, or the probability of more extreme values of family-level statistics. We assumed the standardized PRS of relatives follows a multivariate normal distribution with mean 0, variance 1 and correlation between pairs of relatives equal to twice their kinship coefficient (9). We assumed the PRS only has direct effects on the phenotype and there are no shared environmental effects. We also assumed a rare disease, such that the probability of being unaffected given the PRS can be set to 1, i.e. unaffected subjects do not differ from subjects with unknown phenotype. Under these assumptions, using the Bayes theorem we can express the joint PRS distribution of $n$ relatives in a family where relatives 1 to $k$ are affected and relatives $k$ + 1 to $n$ are unaffected as follows:

$$f\left( g_{1},\ldots,g_{n} | Y_{1}=,\ldots,Y_{k}=1,Y_{k+1},\ldots,Y_{n}=0 \right)=\frac{{{P\left[ Y_{1}=1|g_{1}=0 \right]}^{k}RR}^{g_{1}+\ldots+g_{k}}f_{0,\Sigma}(g_{1},\ldots,g_{n})}{P\left[ Y_{1}=,\ldots,Y_{k}=1,Y_{k+1},\ldots,Y_{n}=0 \right]}$$

where $Y_{i}$ is the affection status (1 = affected, 0 = unaffected), $g_{i}$ is the PRS value for subject $i$, $RR$ represents the relative risk of disease in unrelated individuals for one standard deviation increase in PRS and $f_{0,\Sigma}$ represents the density of the multivariate normal distribution with mean 0 and variance-covariance matrix $S$ which in the present case is twice the kinship matrix. We could not find estimates of SZ and BD PRS effects expressed as $RR$ for one standard deviation increase in PRS in the literature. Instead, we computed the mean PRS in cases and NAARs for selected family structures from the Eastern Quebec Schizophrenia and Bipolar Disorder Kindred~~s~~ cohort using values of $RR$ from 1.3 to 1.5 and assessed the agreement of these values with empirical means of the PRS over the entire sample of cases and NAARs.

The complexity of the computation of the denominator grows exponentially with $n$, so we turn to a Metropolis-Hastings algorithm (10) to sample from the above distribution. We used as proposal distribution at step $t$ the multivariate normal distribution $N(g^{\left( t-1 \right)},0.3I_{n})$ where $g^{\left( t-1 \right)}$ is the PRS vector at step $t-1$ and $I_{n}$ is the identity matrix of dimension $n\times n$. The variance of 0.3 was chosen to ensure good mixing of the sampler for $RR$ between 1.3 and 1.5 with $n$ up to 12 based on inspection of the traces of the sampled values of $g^{\left( t \right)}$. The traces also revealed rapid convergence to the target distribution. From runs of 5,000,000 iterations, we discarded the first 100,000 as burn-in period and sampled 1/5 of the following iterations, resulting in Markov chain Monte Carlo (MCMC) samples of 980,000 observations. We computed exact probabilities that all individual PRS are below a constant with $n=3$ and obtained very close agreement with the MCMC estimate (e.g. 0.3591 for an exact value of 0.3597).

#### 1.5 Clinical validation: Extended phenotypes and symptom dimensions

*Global functioning* was evaluated using the Global Assessment Scale (11) during acute episodes. The *age of disorder onset* was defined as the age at the first definitive or probable episode meeting the DSM-III-R or DSM-IV criteria. *Symptoms dimensions* were assessed by means of the 82 items of the Comprehensive Assessment of Symptoms and History (CASH) instrument (11-13). The lifetime presence and severity of psychotic, manic, and depressive symptoms was evaluated in the lifetime acute episodes. From the multiple sources of information drawn from the defined above lifetime best-estimate procedure, each symptom was rated on a six-point scale, with each point corresponding to an operational definition of severity tailored to the specific symptom: 0 (none), 1 (questionable), 2 (mild), 3 (moderate), 4 (marked), and 5 (severe). This assessment was applied to each of the 82 items of the Comprehensive Assessment of Symptoms and History (CASH DSM-III-R instrument) (N = 85) (11, 12) and SCID DSM IV instrument (N = 288) (14). The original CASH instrument includes 12 symptom dimensions: delusions 15 items), hallucinations (6 items), bizarre behavior (5 items), anhedonia (5 items), apathy (4 items), affective flattening (8 items), thought disorganization (7 items), alogia (4 items), depression (9 items), and mania (8 items). Two domains from the original CASH instrument were excluded from our analysis: *Catatonia* (6 items) due to limited between-subject variability (15), and the *Other Symptoms* category (5 items) (sparse data with > 60% zero values across component items). For a particular dimension, symptom scores were averaged across items within each dimension to produce composite scores. The items defining a dimension are provided in Supplementary Table 1.

To confirm the three-dimensional structure of the CASH in our sample, we performed a principal component analysis (PCA) on the 10 symptom dimension scores in affected family members having complete data (N = 334). Thirty-nine individuals (11.7%) were excluded from the PCA due to complete absence of symptom data. Comparison of these 39 to the 334 others revealed no differences that would affect sample representativeness. Data suitability was confirmed (Bartlett's test p < 0.001; KMO = 0.806). Variables were z-standardized before extraction. Component retention was based on Kaiser criterion (eigenvalue > 1), scree plot and cumulative variance ≥ 70%. The PCA results (Supplementary Figure 1) supported a strong three-component structure explaining 70.5% of the total variance: a schizophrenia component (40.9%), a mania component (17.9%) and a depression component (11.8%).

The *schizophrenia* component yielded high loadings on positive symptoms (delusions (0.709), hallucinations (0.729), bizarre behavior (0.708)) and on negative symptoms (apathy (0.758), alogia (0.729), affective flattening (0.719), anhedonia (0.699)). The *mania* component had two symptomatic poles: high positive loadings for mania (0.848), disorganization (0.578), and bizarre behavior (0.439) and negative loadings for anhedonia (-0.434), affective flattening (-0.392) and apathy (-0.258). The *depression* component was defined solely by the depression symptom (0.836). Individual scores were extracted on each component to characterize a patient in subsequent analyses (see Supplementary Figure 1 for loadings).

### 2 Supplementary Tables

#### Supplementary Table 1 – Mean (SD) raw symptom scores for each symptom dimension of the CASH instrument.

|  | **Mean** | **SD** | **Range** |
| --- | --- | --- | --- |
| Delusions | 0.73 | 0.60 | [0 - 3.20] |
| Hallucinations | 0.88 | 0.84 | [0 - 4.33] |
| Bizarre behavior | 1.07 | 0.90 | [0 - 3.75] |
| Anhedonia | 1.77 | 1.36 | [0 - 5.00] |
| Apathy | 2.13 | 1.37 | [0 - 5.00] |
| Catatonia | 0.18 | 0.32 | [0 - 1.60] |
| Affective flattening | 0.93 | 1.00 | [0 - 5.00] |
| Thought disorganization | 0.99 | 0.87 | [0 - 3.57] |
| Alogia | 0.65 | 0.73 | [0 - 3.50] |
| Depression | 2.34 | 1.04 | [0 - 4.78] |
| Mania | 1.77 | 1.40 | [0 - 4.62] |
| Other symptoms | 1.06 | 0.89 | [0 - 4.00] |

Symptom dimensions were assessed using the 82-item Comprehensive Assessment of Symptoms and History (CASH) DSM-III-R instrument. For each dimension, symptom scores were averaged across items to produce composite scores. The reported range corresponds to the observed minimum and maximum composite scores. These composite scores were then analyzed using a principal component analysis (PCA) (see Supplementary Figure 1). The Catatonia domain was excluded from the PCA because of limited between-subject variability, while the Other Symptoms domain was excluded due to sparse item distributions, with more than 60% zero values across the component items.

PCA, principal component analysis; SD, standard deviation.

#### Supplementary Table 2 – Number of cases and controls depending on the reference group used in the analysis.

| **Phenotype** | **Reference** | **Case (n)** | **Control (n)** |
| --- | --- | --- | --- |
| NAARs (0 dx) | CaG | 320 | 1884 |
| NAARs (≥ 1dx) | NAARs (0 dx) | 140 | 320 |
| NAARs (≥ 1dx) | CaG | 140 | 1884 |
| BD broad | All NAARs | 208 | 460 |
| BD narrow | All NAARs | 127 | 460 |
| BD broad | NAARs (0 dx) | 208 | 320 |
| BD narrow | NAARs (0 dx) | 127 | 320 |
| BD broad | CaG | 208 | 1884 |
| BD narrow | CaG | 127 | 1884 |
| SZ + BD + SAD broad | All NAARs | 373 | 460 |
| SZ + BD + SAD narrow | All NAARs | 281 | 460 |
| SZ + BD + SAD broad | NAARs (0 dx) | 373 | 320 |
| SZ + BD + SAD narrow | NAARs (0 dx) | 281 | 320 |
| SZ + BD + SAD broad | CaG | 373 | 1884 |
| SZ + BD + SAD narrow | CaG | 281 | 1884 |
| SZ broad | All NAARs | 128 | 460 |
| SZ narrow | All NAARs | 119 | 460 |
| SZ broad | NAARs (0 dx) | 128 | 320 |
| SZ narrow | NAARs (0 dx) | 119 | 320 |
| SZ broad | CaG | 128 | 1884 |
| SZ narrow | CaG | 119 | 1884 |

NAARs (0 dx) refers to non-affected adult relatives without other non-mood non psychotic DSM-III-R or DSM-IV diagnosis. NAARs (≥ 1dx) refers to non-affected adult relatives presenting at least one other non-mood non psychotic DSM-III-R or DSM-IV diagnoses. BD narrow includes BD-I only, whereas BD broad includes BD-I, BD-II and recurrent major depression. SZ narrow is restricted to SZ, whereas SZ broad includes SZ narrow plus schizophreniform disorder and schizotypal personality. The broad SZ + BD + SAD phenotype comprises BD broad, SZ broad, and SAD. The narrow SZ + BD + SAD phenotype comprises SZ narrow, BD narrow, and SAD.

NAARs, non-affected adult relatives; CaG*,* CARTaGENE*;* BD, bipolar disorder; SZ, schizophrenia; SAD, schizoaffective disorder.

#### Supplementary Table 3 – Number of variants and subjects for each data source.

| **Data type** | **Source** | **Subjects (n)** | **Variants (n)** | **Variants post-QC (n)** |
| --- | --- | --- | --- | --- |
| Genotyping | OmniExpress | 506 | 651,692 | 600,720 |
|  | GSA | 611 | 691,719 | 470,161 |
| WGS | Eastern Quebec cohort | 464 | 79,000,100 ^a^ | 36,096,233 |
|  | CARTaGENE | 1,884 | 79,000,100 ^a^ | 36,096,233 |

^a^ Contains 3,741,006 variants from chromosome X and Y.

#### Supplementary Table 4 **–** Number of cases (broad phenotypes) and controls depending on the affected parent and the reference group used in the analysis.

| **Phenotype** | **Affected parent** | **Reference** | **Case (n)** | **Control (n)** |
| --- | --- | --- | --- | --- |
| BD broad | Mother | All NAARs | 25 | 460 |
| BD broad | Father | All NAARs | 7 | 460 |
| BD broad | None | All NAARs | 176 | 460 |
| BD broad | Father or Mother | All NAARs | 32 | 460 |
| BD broad | Mother | NAARs (0 dx) | 25 | 320 |
| BD broad | Father | NAARs (0 dx) | 8 | 320 |
| BD broad | None | NAARs (0 dx) | 176 | 320 |
| BD broad | Father or Mother | NAARs (0 dx) | 33 | 320 |
| BD broad | Mother | CaG | 25 | 1884 |
| BD broad | Father | CaG | 8 | 1884 |
| BD broad | None | CaG | 176 | 1884 |
| BD broad | Father or Mother | CaG | 33 | 1884 |
| SZ + BD + SAD broad | Mother | All NAARs | 62 | 460 |
| SZ + BD + SAD broad | Father | All NAARs | 23 | 460 |
| SZ + BD + SAD broad | None | All NAARs | 288 | 460 |
| SZ + BD + SAD broad | Father or Mother | All NAARs | 85 | 460 |
| SZ + BD + SAD broad | Mother | NAARs (0 dx) | 62 | 320 |
| SZ + BD + SAD broad | Father | NAARs (0 dx) | 24 | 320 |
| SZ + BD + SAD broad | None | NAARs (0 dx) | 288 | 320 |
| SZ + BD + SAD broad | Father or Mother | NAARs (0 dx) | 86 | 320 |
| SZ + BD + SAD broad | Mother | CaG | 62 | 1884 |
| SZ + BD + SAD broad | Father | CaG | 24 | 1884 |
| SZ + BD + SAD broad | None | CaG | 288 | 1884 |
| SZ + BD + SAD broad | Father or Mother | CaG | 86 | 1884 |
| SZ broad | Mother | All NAARs | 6 | 460 |
| SZ broad | Father | All NAARs | 3 | 460 |
| SZ broad | None | All NAARs | 119 | 460 |
| SZ broad | Father or Mother | All NAARs | 9 | 460 |
| SZ broad | Mother | NAARs (0 dx) | 6 | 320 |
| SZ broad | Father | NAARs (0 dx) | 3 | 320 |
| SZ broad | None | NAARs (0 dx) | 119 | 320 |
| SZ broad | Father or Mother | NAARs (0 dx) | 9 | 320 |
| SZ broad | Mother | CaG | 6 | 1884 |
| SZ broad | Father | CaG | 3 | 1884 |
| SZ broad | None | CaG | 119 | 1884 |
| SZ broad | Father or Mother | CaG | 9 | 1884 |

NAARs (0 dx) refers to non-affected adult relatives without other non-mood non psychotic DSM-III-R or DSM-IV diagnosis. NAARs (≥ 1dx) refers to non-affected adult relatives presenting at least one other non-mood non psychotic DSM-III-R or DSM-IV diagnoses. BD narrow includes BD-I only, whereas BD broad includes BD-I, BD-II and recurrent major depression. SZ narrow is restricted to SZ, whereas SZ broad includes SZ narrow plus schizophreniform disorder and schizotypal personality. The broad SZ + BD + SAD phenotype comprises BD broad, SZ broad, and SAD. The narrow SZ + BD + SAD phenotype comprises SZ narrow, BD narrow, and SAD.

NAARs, non-affected adult relatives; CaG*,* CARTaGENE*;* BD, bipolar disorder; SZ, schizophrenia; SAD, schizoaffective disorder.

#### Supplementary Table 5 **–** BD PRS mean and pooled standard deviation within the 48 families for the broad SZ + BD + SAD phenotype.

| Cases | | NAARs | | Pooled BD PRS SD |
| --- | --- | --- | --- | --- |
| N | BD PRS mean | N | BD PRS mean |  |
| 2 | -0.87 | 1 | -0.27 | 0.198 |
| 5 | 0.34 | 5 | 0.47 | 0.291 |
| 3 | 0.81 | 6 | 1.002 | 0.414 |
| 4 | 0.438 | 2 | -0.2 | 0.46 |
| **3** | **1.24 ^B^** | **5** | **0.264** | **0.492** |
| 2 | 0.95 | 2 | -0.015 | 0.531 |
| **3** | **-0.277 ^A^** | **5** | **-0.658** | **0.541** |
| 12 | 1.047 | 7 | 0.901 | 0.604 |
| 5 | 2.638 | 0 | NA | 0.613 |
| 5 | 0.662 | 7 | 0.146 | 0.624 |
| 6 | 0.298 | 15 | 0.187 | 0.674 |
| 8 | -0.195 | 9 | -0.2 | 0.679 |
| 4 | 1.675 | 5 | 0.718 | 0.687 |
| 10 | 0.351 | 9 | 0.167 | 0.694 |
| 4 | 1.005 | 4 | 1.592 | 0.715 |
| 6 | 1.632 | 8 | 0.948 | 0.724 |
| 11 | 0.647 | 13 | 0.27 | 0.724 |
| 7 | 0.867 | 12 | 0.27 | 0.735 |
| 4 | 0.93 | 10 | -0.037 | 0.748 |
| 4 | 0.915 | 3 | 1.24 | 0.749 |
| 5 | 1.166 | 5 | 0.536 | 0.749 |
| 8 | 1.224 | 7 | 1.036 | 0.751 |
| 8 | 1.156 | 10 | -0.5 | 0.783 |
| 9 | 1.654 | 10 | 0.825 | 0.784 |
| 7 | 0.517 | 10 | 0.71 | 0.794 |
| 19 | 1.003 | 39 | 0.364 | 0.804 |
| 5 | 1.13 | 3 | 1.263 | 0.806 |
| 10 | 1.151 | 7 | 0.917 | 0.809 |
| 11 | 0.281 | 19 | -0.279 | 0.834 |
| 13 | -0.162 | 35 | -0.526 | 0.852 |
| 10 | 1.554 | 22 | 0.082 | 0.853 |
| 12 | 0.855 | 13 | 0.142 | 0.856 |
| 12 | 0.29 | 12 | -0.41 | 0.893 |
| 6 | 0.51 | 3 | -0.153 | 0.897 |
| 6 | 1.108 | 4 | 0.6 | 0.907 |
| 13 | 0.644 | 18 | 0.494 | 0.926 |
| 7 | 1.09 | 3 | 1.503 | 0.931 |
| 7 | 0.32 | 8 | -0.33 | 0.938 |
| 6 | 1.715 | 6 | 0.543 | 0.956 |
| 17 | 0.373 | 13 | -0.232 | 0.973 |
| 5 | 0.71 | 4 | 0.145 | 1.011 |
| 20 | 0.814 | 33 | 0.209 | 1.021 |
| 8 | 0.532 | 7 | 0.027 | 1.023 |
| 9 | 1.242 | 5 | 0.158 | 1.048 |
| 13 | 0.541 | 17 | 0.1 | 1.063 |
| 3 | 0.907 | 0 | NA | 1.082 |
| 6 | 1.978 | 8 | 1.886 | 1.125 |
| 11 | 0.766 | 10 | 0.145 | 1.177 |

Bold lines are presented in Figure 2A and Figure 2B, as indexed by exponent letters. The broad SZ + BD + SAD phenotype comprises BD broad, SZ broad, and SAD.

$$Pooled PRS SD= \sqrt{\frac{\left( n_{cases}-1 \right)*{Var}_{cases}+\left( n_{NAARs}-1 \right)*{Var}_{NAARs}}{\left( n_{cases}+n_{NAARs}-2 \right)}}$$

BD, bipolar disorder; SZ, schizophrenia; SAD, schizoaffective disorder; NAARs, non-affected adult relative; PRS, polygenic risk score; NAARs, non-affected adult relatives; SD, standard deviation.

#### Supplementary Table 6 **–** SZ PRS mean and pooled standard deviation within the 48 families for the broad SZ + BD + SAD phenotype.

| Cases | | NAARs | | Pooled SZ PRS SD |
| --- | --- | --- | --- | --- |
| N | SZ PRS mean | N | SZ PRS mean |  |
| 4 | 0.81 | 5 | 0.44 | 0.308 |
| **4** | **1.005 ^B^** | **3** | **0.53** | **0.372** |
| 6 | 0.568 | 4 | 0.42 | 0.415 |
| 6 | 0.532 | 15 | 0.062 | 0.477 |
| 4 | 0.828 | 2 | 0.825 | 0.533 |
| 4 | 0.452 | 10 | 0.338 | 0.537 |
| 6 | 1.052 | 6 | 0.308 | 0.542 |
| 5 | 0.262 | 4 | -0.25 | 0.543 |
| 3 | 1.307 | 6 | 0.948 | 0.55 |
| 3 | 0.153 | 5 | -0.192 | 0.554 |
| 5 | 1.684 | 0 | NA | 0.577 |
| **4** | **-0.873 ^A^** | **4** | **-0.302** | **0.577** |
| 7 | 1.599 | 3 | 1.337 | 0.634 |
| 13 | 0.271 | 18 | 0.346 | 0.641 |
| 6 | 0.197 | 3 | -0.093 | 0.66 |
| 6 | 1.31 | 8 | 0.286 | 0.663 |
| 5 | 0.476 | 7 | -0.151 | 0.664 |
| 10 | 0.626 | 7 | 0.584 | 0.671 |
| 10 | 0.211 | 9 | 0.419 | 0.672 |
| 6 | 1.567 | 8 | 1.102 | 0.692 |
| 5 | 1.242 | 3 | 0.457 | 0.716 |
| 3 | 0.697 | 5 | 0.124 | 0.722 |
| 8 | 1.155 | 7 | 0.847 | 0.744 |
| 9 | 0.441 | 5 | 0.286 | 0.772 |
| 8 | 0.45 | 7 | 0.627 | 0.775 |
| 5 | 0.668 | 5 | 0.512 | 0.775 |
| 17 | 0.824 | 13 | 0.192 | 0.784 |
| 7 | 0.039 | 8 | -0.161 | 0.785 |
| 12 | 0.621 | 7 | 0.471 | 0.815 |
| 13 | 0.549 | 35 | 0.24 | 0.838 |
| 8 | 0.312 | 9 | 0.031 | 0.858 |
| 7 | 0.639 | 10 | 0.467 | 0.877 |
| 11 | 1.621 | 13 | 1.455 | 0.905 |
| 5 | 0.868 | 5 | -0.156 | 0.939 |
| 2 | -0.365 | 1 | 1.43 | 0.94 |
| 20 | 0.93 | 33 | 0.383 | 0.941 |
| 8 | 1.29 | 10 | 0.642 | 0.945 |
| 7 | 2.09 | 12 | 1.701 | 0.95 |
| 12 | 0.79 | 13 | -0.016 | 0.972 |
| 11 | 1.215 | 10 | 0.615 | 0.979 |
| 2 | 1.305 | 2 | 1.65 | 0.987 |
| 13 | 0.739 | 17 | 0.655 | 1.001 |
| 9 | 1.906 | 10 | 0.943 | 1.009 |
| 11 | 0.803 | 19 | 0.723 | 1.01 |
| 19 | 0.601 | 39 | 0.207 | 1.023 |
| 12 | 1.529 | 12 | 1.025 | 1.026 |
| 10 | 0.867 | 22 | 0.211 | 1.072 |
| 3 | 0.883 | 0 | NA | 1.22 |

Bold lines are presented in Figure 2A and Figure 2B, as indexed by exponent letters. The broad SZ + BD + SAD phenotype comprises BD broad, SZ broad, and SAD.

$Pooled PRS SD= \sqrt{\frac{\left( n_{cases}-1 \right)*{Var}_{cases}+\left( n_{NAARs}-1 \right)*{Var}_{NAARs}}{\left( n_{cases}+n_{NAARs}-2 \right)}}$«

BD, bipolar disorder; SZ, schizophrenia; SAD, schizoaffective disorder; NAARs, non-affected adult relative; PRS, polygenic risk score; NAARs, non-affected adult relatives; SD, standard deviation.

#### Supplementary Table 7 **–** Probability of observing low PRS (defined as a mean PRS among cases < 0) with the broad SZ + BD + SAD phenotype in families from Figure 2A.

| PRS | SD | P[s < SD] ^a^ | RR ^b^ | Expected case PRS | Expected NAARs PRS | Max PRS | P [all PRS < max PRS] | Max case PRS | P [all case PRS < max case PRS] |
| --- | --- | --- | --- | --- | --- | --- | --- | --- | --- |
| BD | 0.54 | 0.30 | 1.3 | 0.54 | 0.43 | 0.19 | 0.028 | 0.04 | 0.070 |
|  |  | 0.36 | 1.4 | 0.74 | 0.62 |  | 0.017 |  | 0.044 |
|  |  | 0.42 | 1.5 | 0.97 | 0.83 |  | 0.0098 |  | 0.024 |
|  |  |  | Emp ^c^ | 1.01 | 0.45 |  |  |  |  |
| SZ | 0.58 | 0.53 | 1.3 | 0.80 | 0.71 | 0.20 | 0.025 | -0.45 | 0.013 |
|  |  | 0.62 | 1.4 | 1.11 | 1.00 |  | 0.011 |  | 0.0063 |
|  |  | 0.68 | 1.5 | 1.37 | 1.26 |  | 0.0048 |  | 0.0031 |
|  |  |  | Emp ^c^ | 0.91 | 0.22 |  |  |  |  |

^a^ Probability that distribution of PRS in family has a standard deviation lower than observed value SD.

^b^ Relative risk in unrelated individuals for one standard deviation increase in PRS.

^c^ Empirical mean PRS in the entire sample (for comparison with expected values).

The broad SZ + BD + SAD phenotype comprises BD broad, SZ broad, and SAD.

PRS, polygenic risk score; SD, standard deviation; RR, relative risk; BD, bipolar disorder; SZ, schizophrenia; SAD, schizoaffective disorder; NAARs, non-affected adult relative.

#### Supplementary Table 8 **–** Probability of observing high PRS (defined as a mean PRS among cases > 1) with the broad SZ + BD + SAD phenotype in families from Figure 2B.

| PRS | SD | P[s < SD] ^a^ | RR ^b^ | Expected case PRS | Expected NAARs PRS | Min case PRS | P [all case PRS > min case PRS] |
| --- | --- | --- | --- | --- | --- | --- | --- |
| BD | 0.49 | 0.26 | 1.3 | 0.65 | 0.49 | 0.68 | 0.29 |
|  |  | 0.31 | 1.4 | 0.92 | 0.71 |  | 0.43 |
|  |  | 0.35 | 1.5 | 1.17 | 0.92 |  | 0.57 |
|  |  |  | Emp ^c^ | 1.01 | 0.45 |  |  |
| SZ | 0.37 | 0.09 | 1.3 | 0.64 | 0.52 | 0.88 | 0.12 |
|  |  | 0.11 | 1.4 | 0.86 | 0.71 |  | 0.19 |
|  |  | 0.13 | 1.5 | 1.08 | 0.91 |  | 0.29 |
|  |  |  | Emp ^c^ | 0.91 | 0.22 |  |  |

^a^ Probability that distribution of PRS in family has a standard deviation lower than observed value SD.

^b^ Relative risk in unrelated individuals for one standard deviation increase in PRS.

^c^ Empirical mean PRS in the entire sample (for comparison with expected values).

This RR value in the simplified model resulted in mean PRS slightly lower than the empirical means over the sample, while the NAARs mean PRS remained higher than the empirical means.

PRS, polygenic risk score; SD, standard deviation; RR, relative risk; BD, bipolar disorder; SZ, schizophrenia; SAD, schizoaffective disorder; NAARs, non-affected adult relative.

#### Supplementary Table 9 – Description of the subsample (N = 334) of affected family members for whom PCA was performed on symptom dimensions, stratified by sex.

|  | **Total N = 334** | **Female N = 188** | **Male N = 146** |
| --- | --- | --- | --- |
| **Phenotype** |  |  |  |
| BD, n (%) | 179 (53.6) | 115 (61.2) | 64 (43.8) |
| SZ, n (%) | 122 (36.5) | 56 (29.8) | 66 (45.2) |
| SAD, n (%) | 33 (9.9) | 17 (9.0) | 16 (11.0) |
| **Age of disorder onset ^a^** (mean ± SD) | 31.86 ± 11.11 | 32.52 ± 11.33 | 31.02 ± 10.80 |
| **Global functioning ^b^** (mean ± SD) | 28.14 ± 10.14 | 28.99 ± 10.55 | 27.05 ± 9.52 |
| **Individual PRS ^c^** (mean ± SD) |  |  |  |
| BD | 0.85 ± 0.95 | 0.84 ± 0.93 | 0.86 ± 0.99 |
| SZ | 0.86 ± 0.97 | 0.83 ± 0.96 | 0.91 ± 0.98 |
| **Familial PRS ^d^** (mean ± SD) |  |  |  |
| BD | 0.83 ± 0.55 | 0.85 ± 0.496 | 0.80 ± 0.60 |
| SZ | 0.85 ± 0.50 | 0.83 ± 0.49 | 0.87 ± 0.52 |
| **Symptom component score from PCA ^e^** (mean ± SD) |  |  |  |
| Schizophrenia component (PC1) | 0.00 ± 1.0 | -0.14 ± 0.93 | 0.18 ± 1.06 |
| Mania component (PC2) | 0.00 ± 1.0 | 0.04 ± 0.98 | -0.06 ± 1.03 |
| Depression component (PC3) | 0.00 ± 1.0 | 0.10 ± 1.02 | -0.12 ± 0.97 |

^a^ Age of disorder onset was defined as the age (years) at the first definitive or probable episode meeting DSM-III-R or DSM-IV criteria.

^b^ Global functioning was assessed using the GAS during acute episodes (1-100, lower scores = poorer functioning). GAS was missing for 38 participants (M = 16, F = 22).

^c^ Individual PRS were standardized (mean = 0, SD = 1) using CaG as reference.

^d^ Familial PRS were calculated by averaging PRS values across affected family members according to the broad SZ + BD + SAD phenotype definition.

^e^ PCA scores were standardized to have a variance of 1.

The subsample (N = 334) comprises affected family members (broad SZ + BD + SAD phenotype definition) having complete symptom data. The broad SZ + BD + SAD phenotype comprises BD broad, SZ broad, and SAD.

PCA, principal component analysis; BD, bipolar disorder; SZ, schizophrenia; SAD, schizoaffective disorder; GAS, global assessment scale; PRS, polygenic risk score; CaG*,* CARTaGENE; SD, standard deviation.

#### Supplementary Table 10 – Associations between familial PRS and extended phenotypes.

|  |  | **Familial BD PRS ^a^** | | | | | | **Familial SZ PRS ^a^** | | | | | |
| --- | --- | --- | --- | --- | --- | --- | --- | --- | --- | --- | --- | --- | --- |
|  |  | Main effect | | | Interaction with sex | | | Main effect | | | Interaction with sex | | |
|  | N | β  [95% CI] | *p* | *q* | β (Int.) [95% CI] | *p* (Int.) | *q* | β  [95% CI] | *p* | *q* | β (Int.) [95% CI] | *p* (Int.) | *q* |
| **Symptom component from PCA** |  |  |  |  |  |  |  |  |  |  |  |  |  |
| Schizophrenia component (PC1) | 334 | -0.12 [-0.43, 0.19] | 0.439 | 0.780 | -0.06 [-0.50, 0.38] | 0.793 | 0.846 | 0.09 [-0.26, 0.45] | 0.617 | 0.846 | 0.58 [0.03, 1.12] | 0.040 | 0.184 |
| Mania component (PC2) | 334 | 0.31 [0.01, 0.60] | 0.046 | 0.184 | 0.64 [0.20, 1.07] | **0.004** | **0.064** | -0.14 [-0.50, 0.20] | 0.412 | 0.780 | 0.70 [0.14, 1.27] | 0.016 | 0.128 |
| Depression component (PC3) | 334 | 0.13 [-0.15, 0.41] | 0.374 | 0.780 | -0.28 [-0.76, 0.19] | 0.243 | 0.778 | -0.08 [-0.43, 0.27] | 0.667 | 0.846 | 0.28 [-0.29, 0.86] | 0.338 | 0.780 |
| **Clinical characteristics** |  |  |  |  |  |  |  |  |  |  |  |  |  |
| Age of disorder onset ^b^ | 334 | 0.43 [-2.42, 3.27] | 0.769 | 0.846 | - | - | - | -0.93 [-4.42, 2.56] | 0.603 | 0.846 | - | - | - |
| Global functioning ^c^ | 296 | -0.16 [-3.00, 2.67] | 0.910 | 0.910 | - | - | - | 0.57 [-2.69, 3.82] | 0.734 | 0.846 | - | - | - |

^a^ Familial PRS were calculated by averaging PRS values across affected family members according to the broad SZ + BD + SAD phenotype definition.

^b^ Age of disorder onset was defined as the age (years) at the first definitive or probable episode meeting DSM-III-R or DSM-IV criteria.

^c^ Global functioning was assessed using the GAS during acute episodes (1-100, lower scores = poorer functioning).

^d^ Individual PRS were standardized (mean = 0, SD = 1) using CaG as reference.

Results from mixed-effects models adjusted for individual PRS ^d^ and random family intercepts, weighted by inverse-variance. β = Main effect of familial PRS (marginal effect across sexes). *p* = *p* value for main effect. β (Int.) = Interaction coefficient (Familial PRS × Sex). *p* (Int.) = *p* value for interaction. *q* = TSBH-adjusted *p* value controlling false discovery rate, with significance defined at 10% and trends at 20%. β coefficients can be interpreted as effect sizes, as principal component scores were standardized to have a variance of 1. 95% CI = 95% confidence interval.

PCA, principal component analysis; BD, bipolar disorder; SZ, schizophrenia; PRS, polygenic risk score; GAS, global assessment scale; CaG*,* CARTaGENE.

#### Supplementary Table 11 – Associations between familial PRS and extended phenotypes: Sex-stratified effects for significant and trending PRS interactions.

|  |  | **Female** | **Male** | **Interaction** | |
| --- | --- | --- | --- | --- | --- |
| **Symptom component from PCA** | **Familial PRS** | Marginal effect [95% CI] | Marginal effect [95% CI] | *p* | *q* ^a^ |
| Mania component (PC2) | BD | -0.07 [-0.43, 0.30] | **0.57 [0.21, 0.93]** | 0.004 | **0.064** |
| Mania component (PC2) | SZ | -0.45 [-0.88, -0.02] | 0.26 [-0.23, 0.74] | 0.016 | 0.128 |
| Schizophrenia component (PC1) | SZ | -0.16 [-0.58, 0.27] | 0.42 [-0.02, 0.90] | 0.040 | 0.184 |

^a^ Multiple-testing correction used the Benjamini-Hochberg method to control false discovery rate (FDR) at 10% (with trends defined at 20%) across a total of 16 tests.

Stratified effects were estimated from marginal predictions. Marginal effects can be interpreted as effect sizes, as principal component scores were standardized to have a variance of 1. Mixed-effects models were adjusted for individual PRS and random family intercepts and weighted by inverse-variance. Interaction *p* values and FDR-adjusted *q* values were used to assess Sex × Familial PRS interactions. A significant Sex × Familial PRS association was observed between the mania component (PC2) and familial BD PRS (trajectory shown in Supplementary Figure 8). Trends were observed for the association between the mania component (PC2) and Sex × Familial SZ PRS (*q* = 0.128) and between the schizophrenia component (PC1) and Sex × Familial SZ PRS (*q* = 0.184).

PCA, principal component analysis; PRS, polygenic risk score; BD, bipolar disorder; SZ, schizophrenia; FDR, false discovery rate; 95% CI, 95% confidence interval.

### 3 Supplementary Figures

#### Supplementary Figure 1 – Principal component analysis results: Standardized loadings of symptom dimensions on the first three principal components.

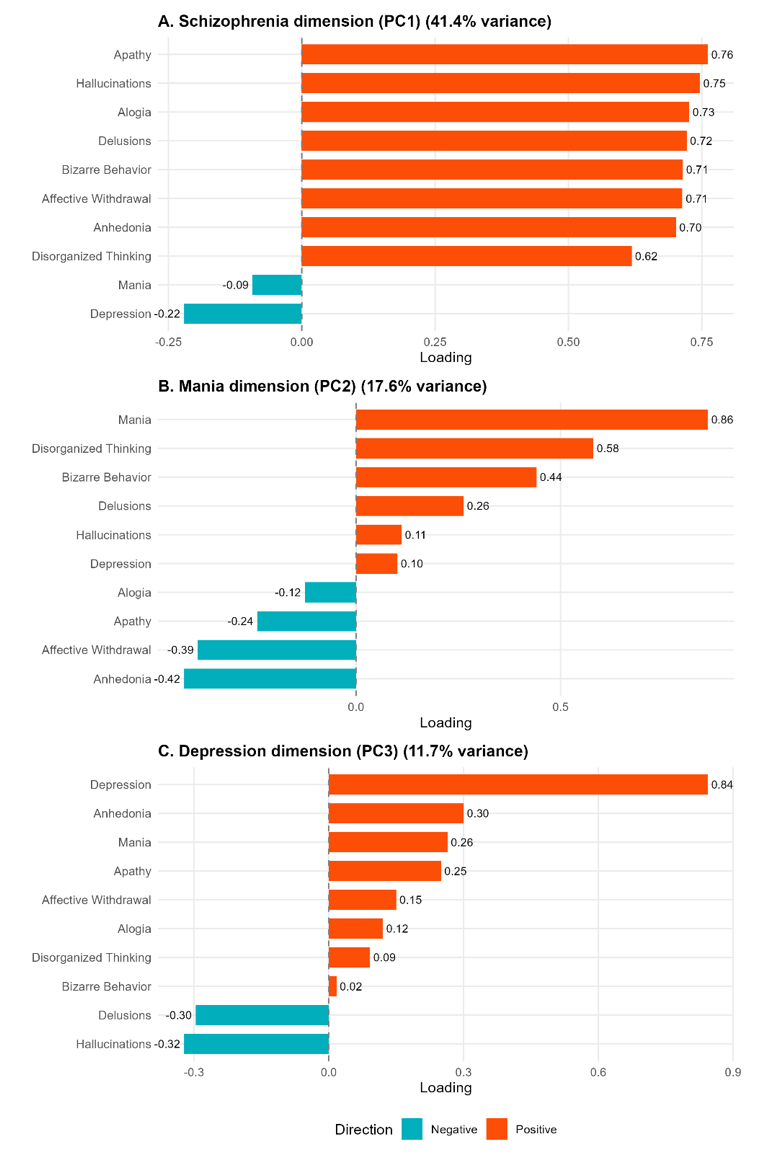

PCA was applied to the mean scores of the 10 symptom dimensions of the Comprehensive Assessment of Symptoms and History (CASH) in affected family members having complete data (N = 334). Variables were z-standardized before extraction and the resulting principal components were orthogonal by construction. See Supplementary Methods 1.4 for details of the PCA procedure. The first three principal components explained 70.5% of the total variance (PC1: schizophrenia component, 40.9%; PC2: mania component, 17.9%; PC3: depression component, 11.8%). Loadings are shown as standardized coefficients (correlations) between each symptom dimension and the corresponding principal component. Positive loadings are displayed in red and negative loadings in blue. As an example, for interpretative purposes, positive and negative scores on the Mania dimension reflect opposing symptom profiles along the mania-depression axis for describing an affected family member.

#### Supplementary Figure 2 – Whole Genome sequencing Quality Control Flowchart.

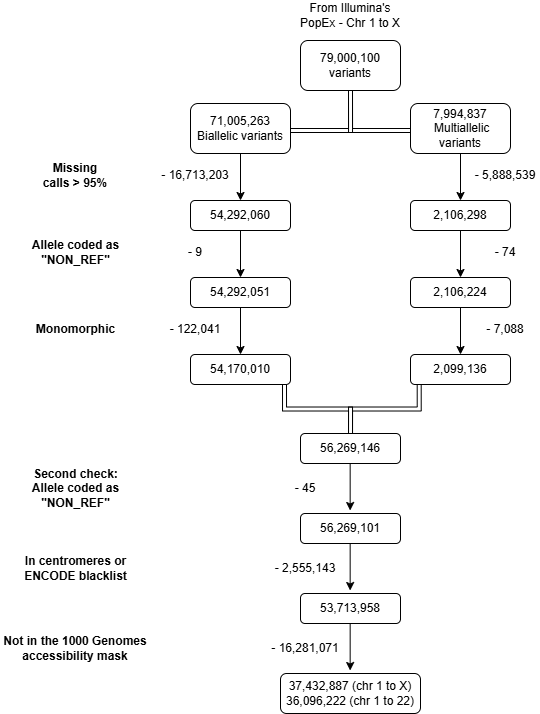

#### Supplementary Figure 3 **–** Principal component analysis (PCA) for the sequencing datasets.

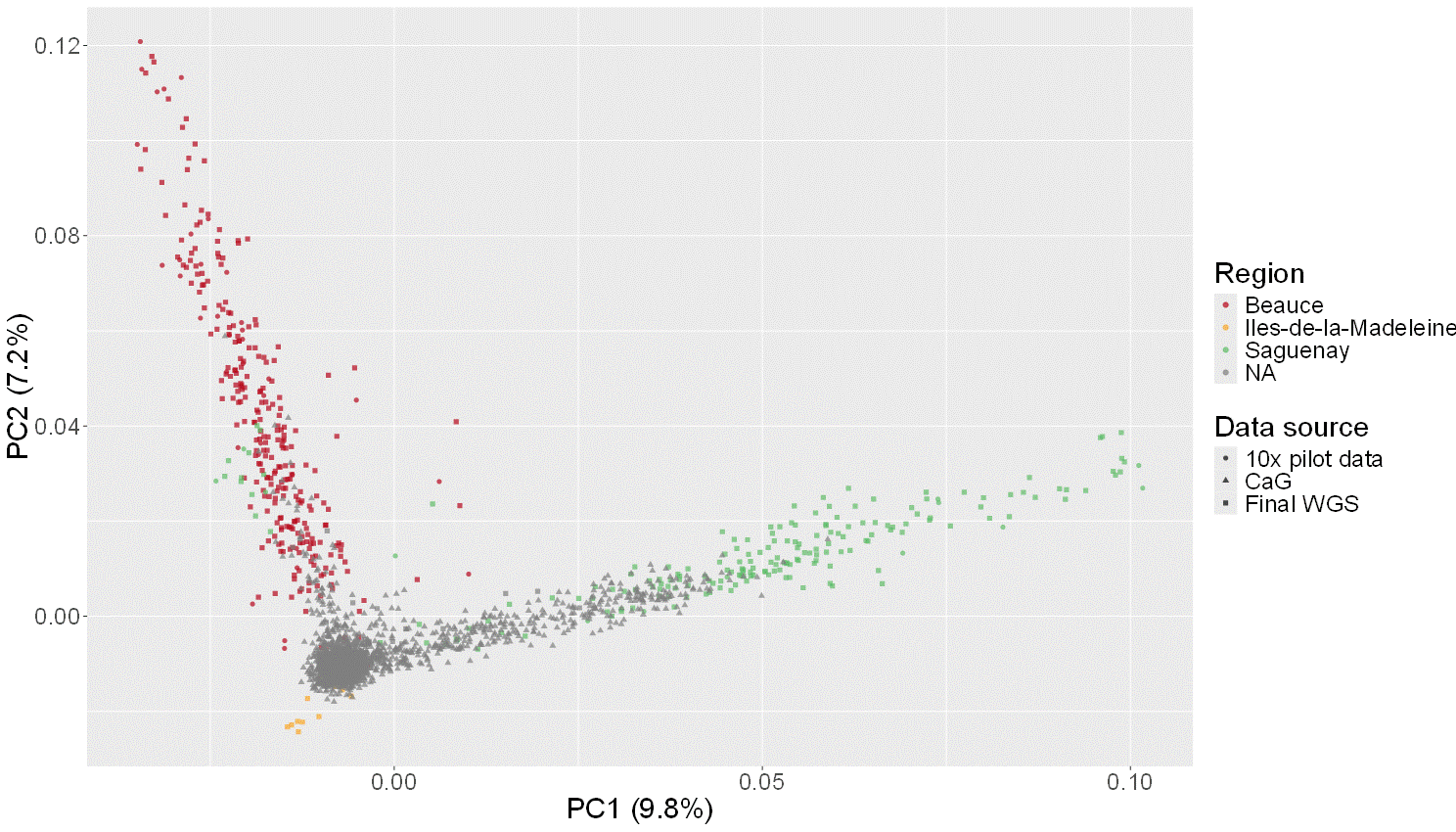

PCA plot to examine the juxtaposition of the 10X pilot data with our final Illumina WGS data and the CARTaGENE (CaG) samples on the principal components.

#### Supplementary Figure 4 **–** Estimated OR and AUC using the continuous PRS by region for the (A) Broad and (B) Narrow phenotypes.

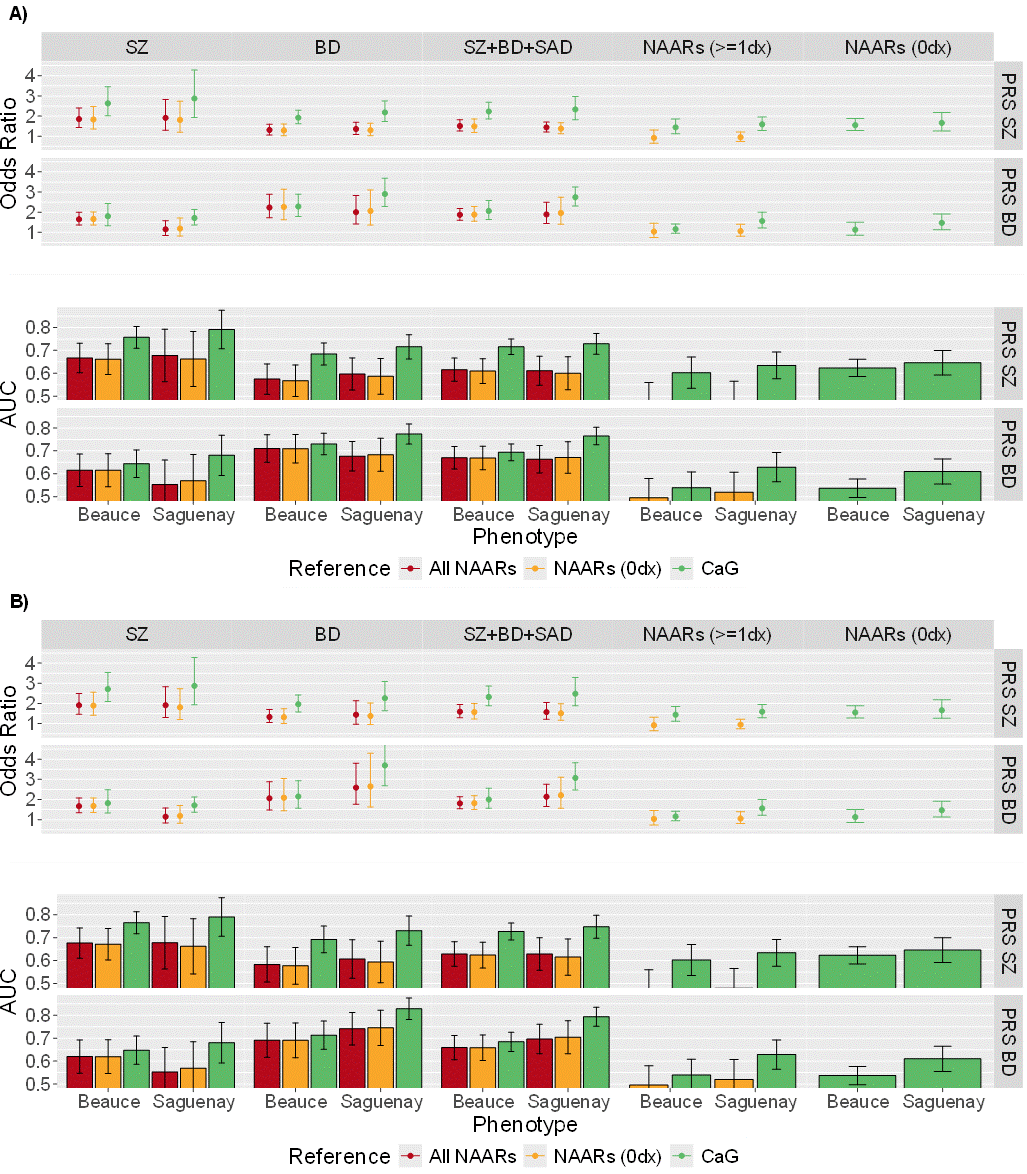

The 57 subjects from the Iles-de-la-Madeleine region (Supplementary Figure 5) were left out of this analysis. NAARs (0 dx) refers to non-affected adult relatives without other non-mood non psychotic DSM-III-R or DSM-IV diagnosis. NAARs (≥ 1dx) refers to non-affected adult relatives presenting at least one other non-mood non psychotic DSM-III-R or DSM-IV diagnoses. BD narrow includes BD-I only, whereas BD broad includes BD-I, BD-II and recurrent major depression. SZ narrow is restricted to SZ, whereas SZ broad includes SZ narrow plus schizophreniform disorder and schizotypal personality. The broad SZ + BD + SAD phenotype comprises BD broad, SZ broad, and SAD. The narrow SZ + BD + SAD phenotype comprises SZ narrow, BD narrow, and SAD.

OR, odds ratio; AUC, Area under the receiver operating curve; PRS, polygenic risk score; NAARs, non-affected adult relatives; BD, bipolar disorder; SZ, schizophrenia; SAD, schizoaffective disorder; CaG, CARTaGENE.

#### Supplementary Figure 5 **–** Estimated OR and AUC using the continuous PRS without the 57 subjects from Iles-de-la-Madeleine.

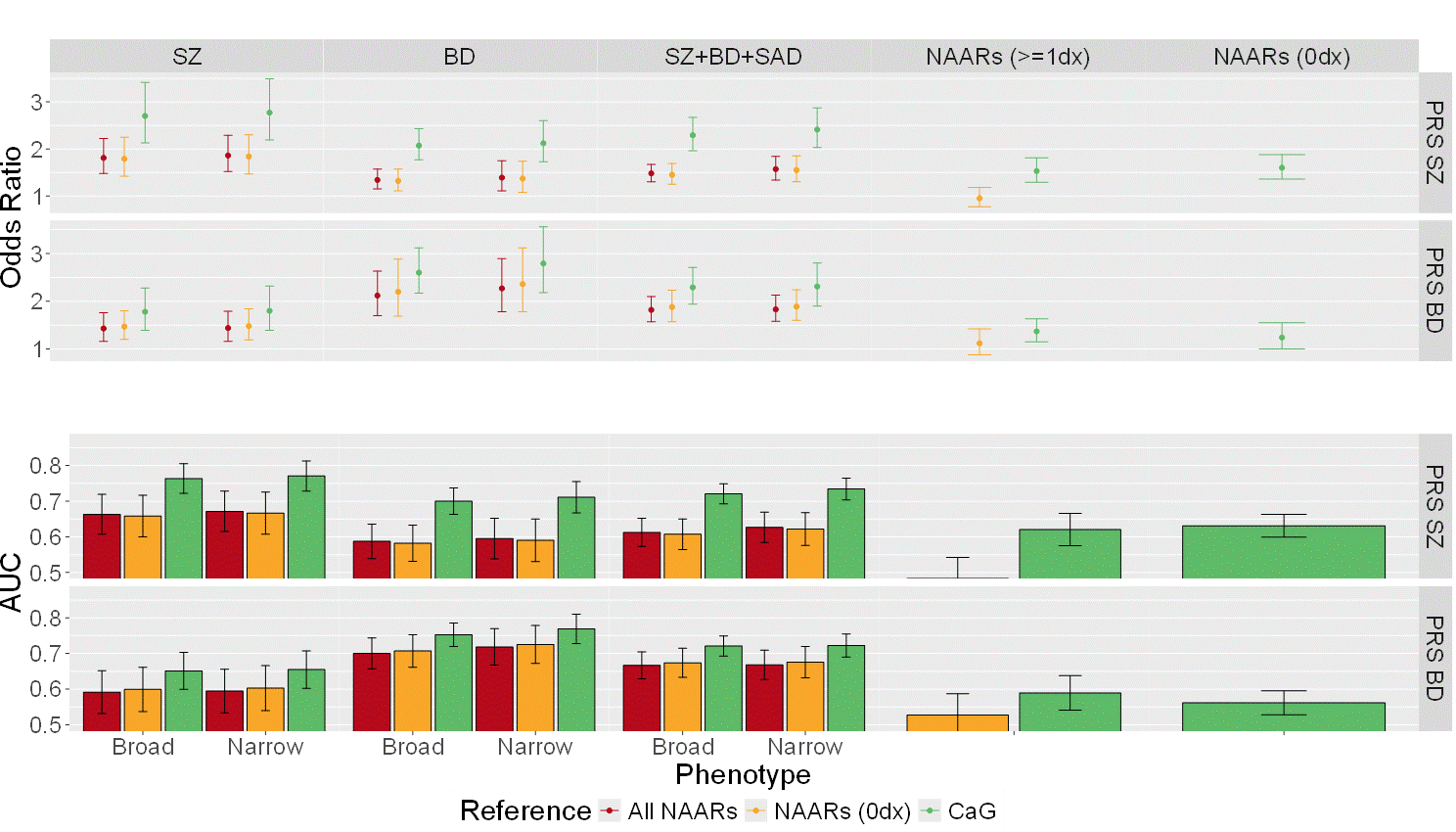

NAARs (0 dx) refers to non-affected adult relatives without other non-mood non psychotic DSM-III-R or DSM-IV diagnosis. NAARs (≥ 1dx) refers to non-affected adult relatives presenting at least one other non-mood non psychotic DSM-III-R or DSM-IV diagnoses. BD narrow includes BD-I only, whereas BD broad includes BD-I, BD-II and recurrent major depression. SZ narrow is restricted to SZ, whereas SZ broad includes SZ narrow plus schizophreniform disorder and schizotypal personality. The broad SZ + BD + SAD phenotype comprises BD broad, SZ broad, and SAD. The narrow SZ + BD + SAD phenotype comprises SZ narrow, BD narrow, and SAD.

OR, odds ratio; AUC, Area under the receiver operating curve; PRS, polygenic risk score; NAARs, non-affected adult relatives*;* BD, bipolar disorder; SZ, schizophrenia; SAD, schizoaffective disorder; CaG*,* CARTaGENE.

#### Supplementary Figure 6 **–** Estimated OR and AUC using the continuous PRS by sex for the (A) Broad and (B) Narrow phenotypes.

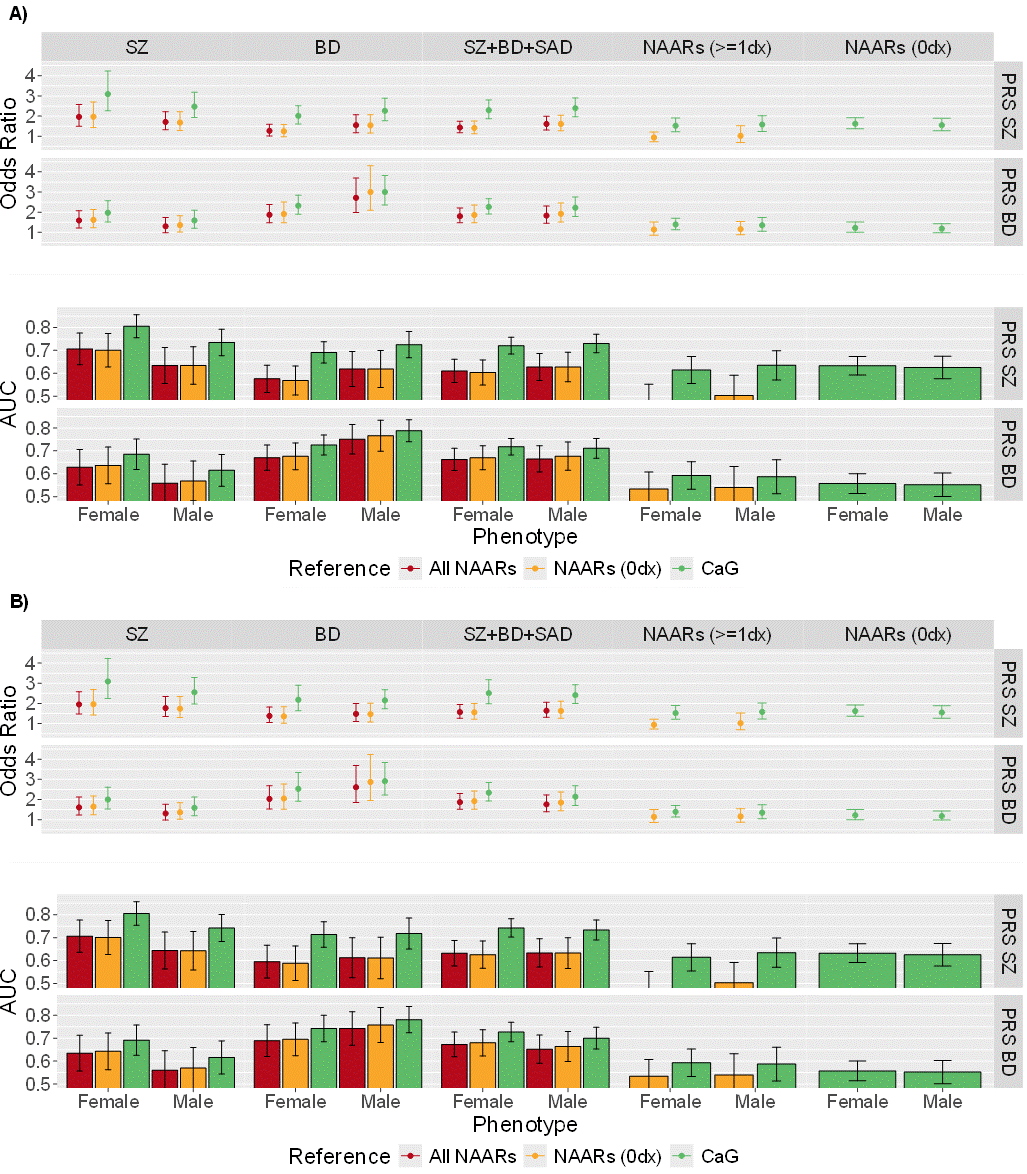

NAARs (0 dx) refers to non-affected adult relatives without other non-mood non psychotic DSM-III-R or DSM-IV diagnosis. NAARs (≥ 1dx) refers to non-affected adult relatives presenting at least one other non-mood non psychotic DSM-III-R or DSM-IV diagnoses. BD narrow includes BD-I only, whereas BD broad includes BD-I, BD-II and recurrent major depression. SZ narrow is restricted to SZ, whereas SZ broad includes SZ narrow plus schizophreniform disorder and schizotypal personality. The broad SZ + BD + SAD phenotype comprises BD broad, SZ broad, and SAD. The narrow SZ + BD + SAD phenotype comprises SZ narrow, BD narrow, and SAD.

OR, odds ratio; AUC, Area under the receiver operating curve; PRS, polygenic risk score; NAARs, non-affected adult relatives*;* BD, bipolar disorder; SZ, schizophrenia; SAD, schizoaffective disorder; CaG*,* CARTaGENE.

#### Supplementary Figure *7* **–** SZ PRS and BD PRS pooled standard deviation (SD) according to the total number NAARs and cases of the broad SZ + BD + SAD phenotype within the 48 families.

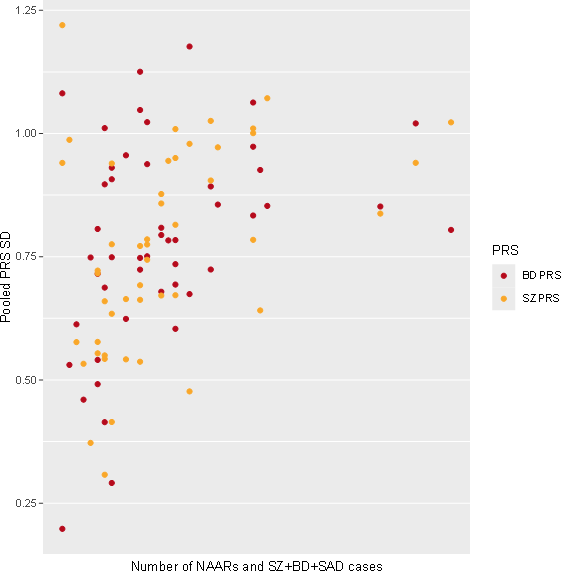

Values are presented in Supplementary Table 6 and Supplementary Table 7. The broad SZ + BD + SAD phenotype comprises BD broad, SZ broad, and SAD.

$$Pooled PRS SD= \sqrt{\frac{\left( n_{cases}-1 \right)*{Var}_{cases}+\left( n_{NAARs}-1 \right)*{Var}_{NAARs}}{\left( n_{cases}+n_{NAARs}-2 \right)}}$$

PRS, polygenic risk score; SD, standard deviation; NAARs, non-affected adult relatives*;* BD, bipolar disorder; SZ, schizophrenia; SAD, schizoaffective disorder.

#### Supplementary Figure 8 – Illustration of the sex-stratified association between familial BD PRS and the mania principal component (PC2) scores.

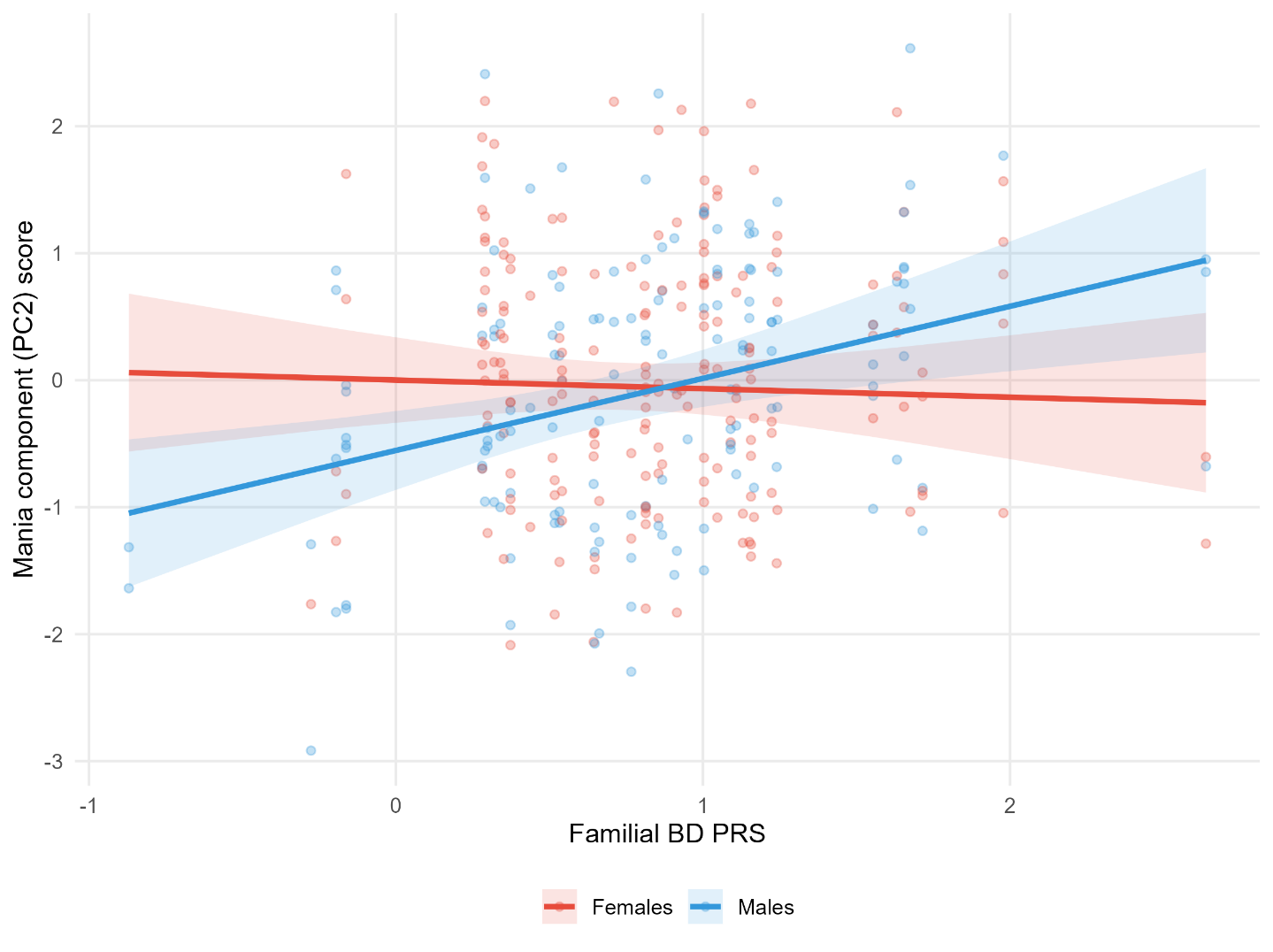

Mania component (PC2) scores as a function of familial BD PRS (Sex × Familial BD PRS interaction: p = 0.004, q = 0.064). Lines represent marginal predictions derived from linear mixed-effects models including familial PRS as the main predictor, adjusted for individual PRS and incorporating random family intercepts; models were weighted by inverse-variance. Sex-stratified marginal effect estimates and confidence intervals are reported in Supplementary Table 11.

PRS, polygenic risk score; BD, bipolar disorder.
